# More than one piece of the puzzle: considering non-clinical factors for personalisation in digital phenotyping

**DOI:** 10.1101/2025.11.18.25340238

**Authors:** Imogen E. Leaning, Linda Schlüter, Imke de Wijs, Mila Brandsen, Mila Roozen, Raj Jagesar, Sarah Tjeerdsma, Anna Tyborowska, Nessa Ikani, Martien J.H. Kas, Christian F. Beckmann, Henricus G. Ruhé, Andre F. Marquand

## Abstract

**Background:** Digital phenotyping is an emerging field that aims to contribute to the clinical care of patients with mental disorders by offering objective, passive behavioural monitoring. This monitoring could be used for applications such as predicting the onset of episodes of mental illness. However, behaviours are often not unique to clinical disorders, and other factors in a person’s life may contribute to their digital phenotyping behavioural pattern.

**Objective:** We aimed to investigate non-clinical factors that may be relevant for personalisation in digital phenotyping, such as the area in which participants live and their regular phone habits, and discussed their implications in a depression relapse case study.

**Methods:** In the MENTALPRECISION study we collected passive smartphone data (phone usage and location behaviours) in a predominantly healthy cohort (n=73) using the Behapp application. We administered a novel questionnaire, the “Smartphone Usage and Lifestyle Questionnaire” (SULQ), to the participants to gather information on their phone usage and lifestyle habits that could impact their digital phenotyping data. We trained a hidden Markov model (HMM) on the smartphone data and developed two types of digital phenotyping measures from the identified hidden behavioural states of the HMM. The “total dwell time” gave the percentage of time each participant spent in each state. The “individual transition probability” was extracted from the HMM itself for each participant, giving their personalised probabilities of transitioning between each of the hidden states. We compared the HMM-generated hidden state sequences and reported events such as holidays and illness. We carried out logistic regression between the digital phenotyping measures and various SULQ measures. We then provide a proof-of-concept for predicting depression relapse in recurrent depression using a HMM and consider the implication of non-clinical factors for this clinical application.

**Results:** Visible differences in behaviour surrounding holidays and illness were observed in the generated hidden state sequences from the MENTALPRECISION study, as well as surrounding the depression relapse in the proof-of-concept. Participants who use another phone in addition to their personal smartphone spent significantly less time in the “socially inactive home time” state (FDR-corrected P=0.03, odds ratio 0.9196, 95% CI 0.8583-0.9808). iOS users spent significantly less time in the “socially active home time” state (FDR-corrected P=0.04, odds ratio 0.9330, 95% CI 0.8804-0.9857) than Android users, and participants reporting a smartphone addiction spent significantly more time in this state (FDR-corrected P=0.009, odds ratio 1.0787, 95% CI 1.0301-1.1272) when compared to participants reporting no smartphone addiction. Relationships between the mean transition probabilities and SULQ measures did not survive multiple comparison correction. We observed decreases in the monthly likelihood surrounding the depression relapse period, providing a possible metric for relapse prediction.

**Conclusions:** When searching for clinically relevant behavioural signals, digital phenotyping researchers should consider additional non-clinical factors that may be contributing to the measured digital signal. Including this additional information could be used to improve personalisation of digital phenotyping models, leading to improved modelling abilities for goals such as depression relapse prediction.

## Introduction

As many areas of healthcare increasingly turn to digital support tools, behavioural monitoring, on the basis of digital devices that can passively collect data such as smartphones, has garnered interest in mental healthcare due to its ability to objectively and unobtrusively quantify behaviour. This is referred to as “digital phenotyping”, which involves quantifying behavioural or physiological markers using digital devices. Digital phenotyping encompasses general behaviours measured using a digital signal, such as periods of travel, and digital-specific behaviours, such as phone usage. Digital phenotyping aims to improve patient monitoring through enabling high temporal resolution behavioural and symptom tracking and has the potential to achieve goals such as the prediction of relapse in people with depression.

Digital phenotyping has several features that make it attractive for clinical applications. Data is collected passively and continuously, meaning that burden is reduced as users do not need to actively participate in data collection. It has a high temporal resolution, meaning that it is in principle suitable for making time-resolved predictions, for example signalling an imminent episode of mental illness. Also, data collection occurs as people go about their daily lives, meaning that it has high ecological validity. There is no risk of recall bias, as measurements are recorded objectively. These proxy measurements of behaviour are referred to as “digital phenotypes”. Digital phenotyping data can be collected using various devices, such as smartwatches (e.g. [1]) and smartrings (e.g. [2]). We focus on smartphones due to their ubiquity in modern society. In order to establish the clinical value of digital phenotyping, many studies have focused on investigating relationships between digital phenotypes and clinical outcomes, such as depressive symptom severity [3-9]. These studies have shown promising results, with digital phenotyping depression prediction models generally achieving moderate performance [10]. However, there are two important factors that limit the clinical utility of these models: first, there has historically been a limited degree of personalisation of the models used to analyse such data, which is important because digital phenotypes are highly individualised. Second, the effect of individual factors such as lifestyle, demographic differences and daily routine are typically not assessed. Yet, this is crucial to ensure that behavioural changes identified using digital phenotyping can be attributed to clinically relevant effects, rather than other factors.

In order to improve prediction models, recent digital phenotyping studies are shifting towards including some level of personalisation. This invariably involves a trade-off in that purely individual models are completely tailored to one person’s data, thereby limiting the size of the training dataset. In contrast, in group-based models, individuals are pooled together for model training to derive a model representative of the group. Whilst from a personalisation perspective group-based models sound undesirable, they allow for larger datasets to be used in training. A middle ground could be used to maximise dataset size whilst allowing for personalisation. For example, Scott et al. [11] allowed for both group and individual parameters in their timeseries models of symptomatic psychosis substates trained using self-report data. Models including personalisation have shown improved performance in studies predicting anhedonia [12], mood episodes [13] and depressive symptom severity [14]. In Langholm et al. [12], within-participant models (including support vector machines, random forest and decision trees) predicting anhedonia scores consistently outperformed between-participant models. In Cho et al. [13], random forest models trained on individual data were compared to group-based models, with personalised models almost always outperforming group models. In Kathan et al. [14], three different approaches were investigated: a transfer learning approach including shared and personalised layers, a group-based model including subject-dependent standardisation, and a subgrouping approach where different models were trained for female and male participants. The transfer learning approach provided the highest performance when predicting following-day depression, with all personalisation model types outperforming the baseline model with no personalisation.

With regard to different factors that affect digital phenotyping data, Kathan et al. [14] also addressed differences in predictive performance of digital phenotyping models between females and males. They identified that baseline model performance was consistently higher for female participants than for male participants, with model personalisation improving fairness. Their study is an example of how personalisation does not just relate to the modelling decisions that should be made to accommodate individuals, but highlights that individual factors should be evaluated as model performance may vary. Further, individual differences in digital phenotypes have been observed for factors such as age and employment status [7,15]. Another study investigated the performance of a general population model for depression prediction in various subgroups created based on gender, age, education, urban/rural environment, sexual orientation and ethnicity [16]. However, model performance was generally low, and not reliably improved by training subgroup models, showing the difficulties of tailoring models to various groups.

Many of these investigated factors have been simple demographic features, however a person’s daily routine is likely shaped by many other factors. Meanwhile, given the range of behavioural symptoms people with mental disorders experience, for example social withdrawal [17], clinically-focused digital phenotyping studies operate under the assumption that the digital behavioural patterns contain clinically-relevant signals. We propose that in order for digital phenotyping to reach its full clinical potential in capturing individual behavioural profiles for personalised prediction, researchers need to account for other possible non-clinical sources of behavioural signals that can influence digital phenotyping patterns. For example, a person’s work, family, lifestyle and adopted phone usage routines are likely to have a strong influence on their digital phenotyping data. A person may have higher levels of phone usage due to needing to use their phone for work, or may spend more time travelling if they live in a rural area with no nearby amenities. These sources of signal conflate the clinical signal expected to be seen in digital behavioural patterns and should be accounted for in the analysis.

For example, Pillai et al. [18] used multi-task anomaly detection to detect a range of rare life events (such as personal, work, and health events) in a non-clinical sample, and in a study of college students, non-clinical factors have been explored [19]. For example, behavioural differences between students’ first versus final terms, such as fewer unique locations visited, were identified. Whilst not explicitly addressing non-clinical factors, in Calvert et al. [20] digital phenotypes are regularly discussed with patients, providing the opportunity for non-clinical factors to be addressed. Without some amount of contextual knowledge, it may be difficult to distinguish between clinically and non-clinically relevant changes in digital phenotyping behavioural profiles, for example the pattern of a person staying at home due to a depression relapse versus staying at home due to the adoption of a work-from-home routine.

Alongside evaluating predominantly non-clinical life events, Pillai et al. [18] demonstrated a time-resolved approach to anomalous behaviour detection. With clinical implementation in mind, digital phenotyping is making efforts towards time-resolved analysis of behavioural patterns, i.e. through using modelling approaches that allow behavioural changes to be identified along a time series (for example in the lead up to a depression relapse). For example, Fujino et al. [21] identified a decrease in mean daily step counts in the two weeks leading up to depression-related medical visits, and Cohen et al. [22] used a combination of passive and active measures to predict changes in depression and anxiety symptoms using anomaly detection methods. Similarly, Cohen et al. and Henson et al. [23,24] used an anomaly detection approach to investigate schizophrenia relapse in populations in the US and India.

In this study, we aimed to further the personalisation of digital phenotyping by investigating the relationship between non-clinical factors and digital phenotypes. Although our approach is widely applicable, to develop our understanding of what factors need to be considered to disentangle clinical signals from other behavioural signals (see Figure 1), we utilised digital phenotyping features derived from a previously validated hidden Markov model (HMM) approach [25]. We trained a group-based HMM on digital phenotyping data collected during the ongoing MENTALPRECISION study using the Behapp smartphone application. This study aims to integrate biology and behaviour in healthy and cross-diagnostic patient cohorts for precision stratification of mental disorders. We included some individual parameters in the HMM to allow for personalisation in the model. We then considered non-clinical factors such as a person’s adopted phone usage habits (e.g. using their phone for work, having a secondary phone) and lifestyle factors (e.g. the area in which they live, their work schedule) using a novel questionnaire we developed, namely the “Smartphone Usage and Lifestyle Questionnaire” (SULQ). We investigated relationships between SULQ responses and HMM-derived digital phenotypes. Next, we supplement our investigations of non-clinical factors in digital phenotyping by showing a proof-of-concept application of digital phenotyping methods to predict relapse in recurrent depression, by identifying behavioural changes in the time series preceding the relapse period using anomaly detection (Figure 2). Our approach involves training a personalised HMM on “baseline” data, that is, periods without a depression relapse, and detecting deviations from this baseline by calculating the likelihood of unseen periods (given the “baseline” HMM). We hypothesised that periods during which a relapse occurred would have a lower likelihood than periods without a relapse, due to the expected deviations in behaviour that could indicate relapse. Finally, considering this depression relapse case study, we discuss how to consolidate clinical and non-clinical behavioural signals for clinical applications.

**Figure 1:**
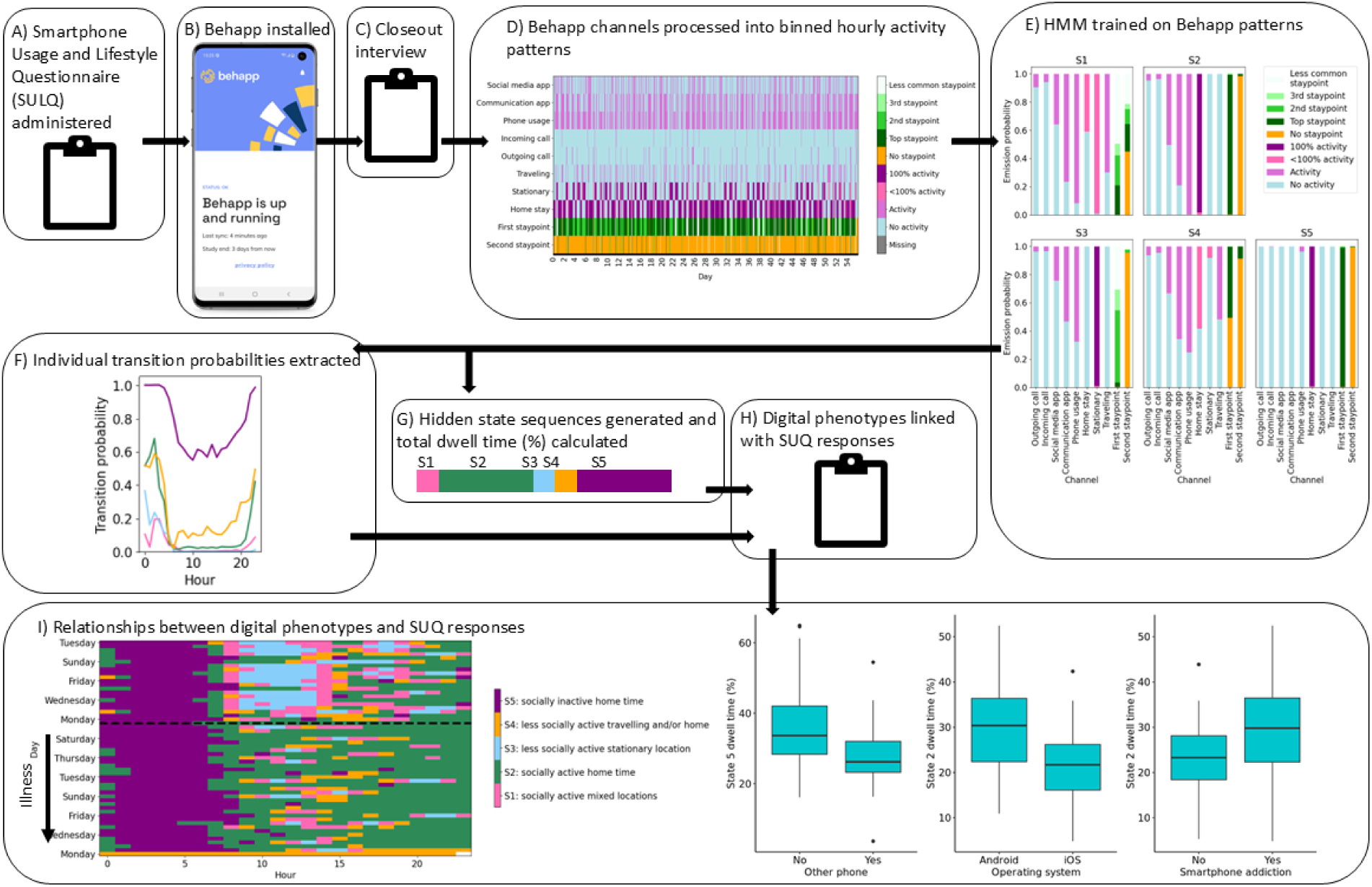
Overview of the Smartphone Usage and Lifestyle Questionnaire (SULQ) investigations. A) Before Behapp participation commenced, the SULQ was administered. B) Behapp data were then recorded for 8 weeks. C) After Behapp participation, a closeout interview was administered, during which participants were asked about any out of the ordinary events that happened during Behapp data collection. D) Behapp data were processed and aggregated into hourly bins. E) A hidden Markov model (HMM) was trained on the digital phenotyping data and the HMM emission probabilities used to interpret the hidden states. F) and G) Digital phenotypes were extracted from the HMM and H) compared to SULQ items. I) Obvious individual case behavioural differences were observed around events such as illness (lefthand side), and significant associations between SULQ measures and total dwell times were observed (righthand side).

**Figure 2:**
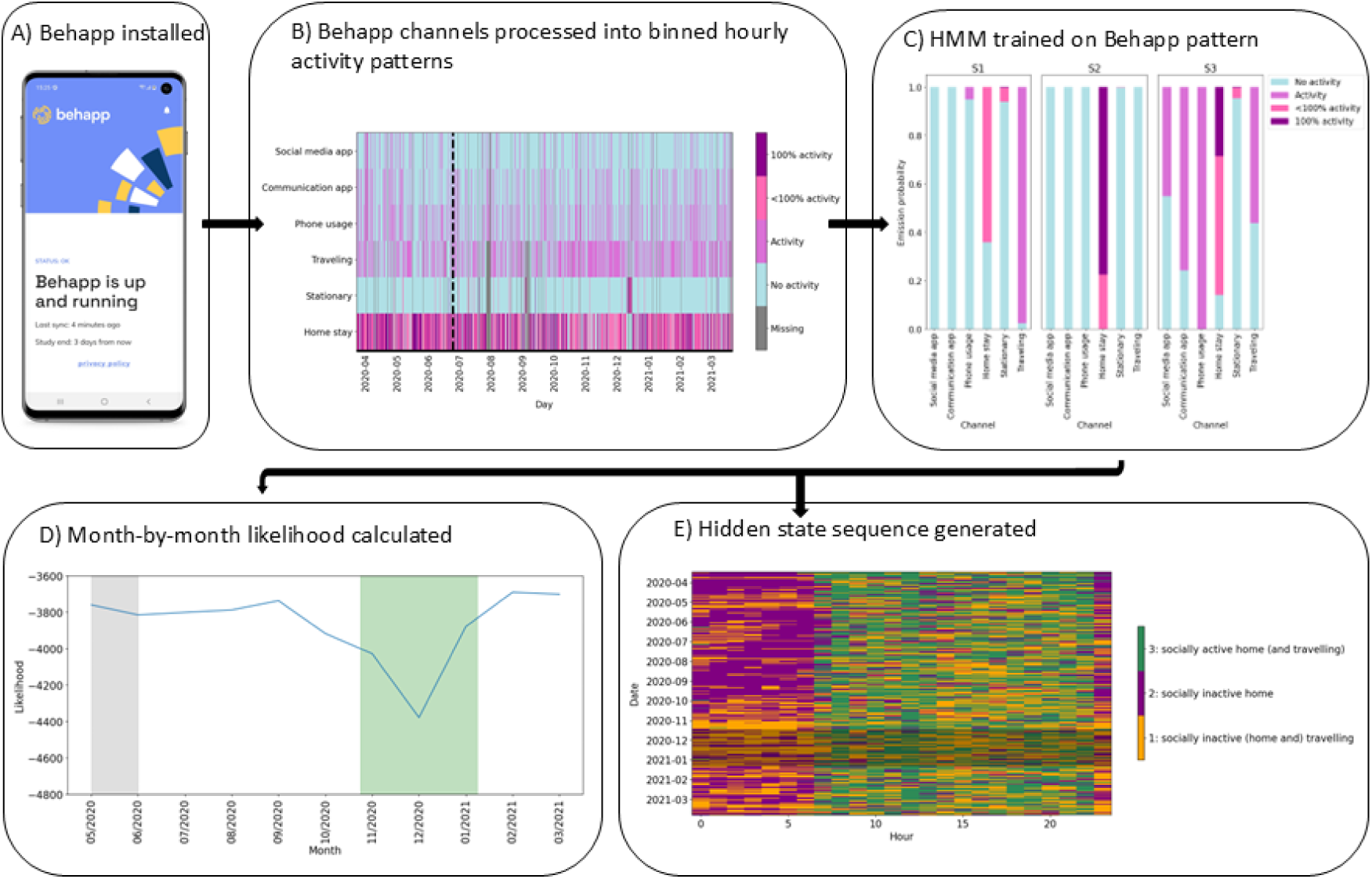
Overview of the relapse prediction method. As in Figure 1, A) Behapp was installed on the participant’s phone, B) the Behapp channels were processed into hourly bins and C) a hidden Markov model (HMM) was trained on the Behapp time series and the HMM emission probabilities used to interpret the hidden states. The steps then deviated: D) the likelihood of each month, given the trained HMM, was calculated, and lower likelihood scores were observed just before and during the months where a relapse was recorded. E) The hidden state sequence was generated to observe behavioural differences around the relapse period.

## Methods

This study consisted of two parts: in part one, non-clinical factors were investigated using the MENTALPRECISION dataset and in part two, a relapse case study was carried out using a participant from the Smartphone-based Monitoring and cognition Modification Against Recurrence of Depression (SMARD) dataset.

### Participants

#### MENTALPRECISION

The MENTALPRECISION study involves acquiring data from healthy controls (HCs) and cross-diagnostic patients in the Nijmegen area in the Netherlands. Eligible HCs were referred to MENTALPRECISION by the Healthy Brain Study (HBS) [26], the Advanced Brain Imaging database on ageing and memory (ABRIM) [27], and the Measuring Integrated Novel Dimensions in Neurodevelopmental and Stress-Related Mental Disorders (MIND-Set) study [28]. All participants from these studies had provided permission to be contacted about participation in future studies. Whilst neuroimaging data were also acquired, these will be reported separately. HCs were also recruited via the standard recruitment procedures at the Donders Centre for Cognitive Neuroimaging, which is also where intake visits and any additional MRI scanning took place for all participants. HCs were required to be between 16-80 years of age.

The cross-diagnostic patient cohort was recruited via the MIND-Set 2 study [28], which was open to patients 16 years of age or older at the Psychiatry department at the Radboudumc in Nijmegen, who were referred to the department due to stress-related (mood, anxiety, addiction and/or mood disorder with somatic co-morbidity), neurodevelopmental (autistic spectrum disorder, attention deficit hyperactivity disorder and/or personality disorders) and/or impulse-regulation disorders. Diagnosis was confirmed during intake. As with the HCs, these participants had provided permission to be contacted about participating in future studies and were asked by their care provider if they would like to participate in MENTALPRECISION. These participants were invited to undergo neuroimaging following the neuroimaging protocols of MIND-Set 1 [28].

For participation in the Behapp portion of the study, participants were required to have and use a smartphone. Participants who did not speak Dutch or English, who had a prior history of significant neurological illness, were pregnant, currently experiencing a psychosis or who were not able/willing to provide informed consent were excluded. Additionally, HCs were excluded if they were currently using medication that targets the brain, had a current disease affecting the brain (e.g. epilepsy) or a neurological disorder of the central nervous system, a prior history of significant psychiatric illness, high alcohol intake per week (fifteen or more units), weekly recreational drug use (once or more per week), used benzodiazepines once or more per week, had malignancies, rare, chronic somatic disorders or first degree relatives with severe psychiatric disorders.

Behapp data were collected for 8 weeks. For inclusion in the current study, we set a generous minimum data availability criteria on the Behapp time series of each participant, to ensure that any participants with major data collection issues would be excluded, requiring at least 75% of datapoints from seven consecutive days to be present in a participant’s time series.

#### SMARD

We include a proof-of-concept analysis using a participant selected from the SMARD study in the Netherlands (selection made on the basis of data quality). The aim of the SMARD study is to investigate recurrent depression in MDD. Participants between the ages of 18 and 65 years, who had previously been diagnosed with MDD, had experienced ≥3 prior depressive episodes and were in stable remission at the start of study participation were recruited [29]. Classification of MDD was made according to the Structured Clinical Interview for Diagnostic and Statistical Manual of Mental Disorders, fourth edition (DSM-IV) (SCID) [30], and stable remission was defined as having a Hamilton Depression Rating Scale score ≤10 for ≥8 weeks [31]. Further information regarding inclusion and exclusion criteria is provided elsewhere [29]. Participants were required to be regular smartphone users, due to the Behapp data collection. Participants downloaded Behapp on their smartphones and passive data collection was carried out for 1.5 years. Every three months, participants underwent clinical interviews (SCID-I and questionnaires), during which any new depressive episodes were identified.

SMARD participants with high data availability have been used previously in our imputation research [32]. However, many of the SMARD participants who had experienced a relapse, who participated during this first phase of data collection (considered to be participation between August 2019 – March 2023 when an earlier Behapp version was predominantly used in SMARD), had high rates of missingness in their Behapp time series, hindering our relapse-specific investigations. Fortunately, one of these participants had very high data availability across channels (2% missing across all channels, 6% missing in GPS channels), and was therefore selected for use in our proof-of-concept investigation.

### Ethical approval and informed consent

MENTALPRECISION and SMARD were each approved by the Medical Ethics Committee for the East of the Netherlands (“METC Oost Nederland”, MENTALPRECISION dossier number: NL82527.091.22, SMARD dossier number: NL60033.091.16). All participants provided informed consent before participating in any part of the MENTALPRECISION or SMARD study. MENTALPRECISION participants received €30 for the Behapp portion of the study plus travel expenses. SMARD participants received compensation of €40 plus travel expenses once they had completed the 1.5 year study. All participants’ data were deidentified and only the authors on the Behapp team (MR, RJ and MK) had access to the raw GPS coordinates.

### Behapp acquisition

Behapp [33] recorded location and general phone usage data for each participant. Phone usage content was not collected, in compliance with the GDPR [34]. Raw GPS locations and their corresponding timestamps were collected for all participants. For Android users, as in previous studies, phone usage data included communication app usage, social media app usage, incoming/outgoing calls and an overall phone usage measure (showing the periods the phone was unlocked). Apps were labelled according to their Google Play Store classification. For each app use or call, the corresponding start and end time was recorded.

Whilst previous Behapp versions were only able to collect data from Android smartphones, the Behapp version used in the MENTALPRECISION study was also able to collect data from iOS smartphones. However, due to iOS restrictions the app usage channels were not available, and the collection of the overall phone usage measure was impacted. As a replacement for the original overall phone usage measure, a new measure was introduced that was derived from the screen status, as this could be measured for both Android and iOS. There were minor differences in the screen status that could be recorded, with possible statuses for Android being “screen off”, “screen on but locked” and “screen on and unlocked”. For iOS, only “screen off” and “screen on and unlocked” could be recorded. Occasionally, a “screen on” status could be recorded but no associated “off” status occurred. To reduce inaccuracies caused by this, Behapp carried out scheduled screen checks and recorded the screen status at this time. The app version used in the SMARD study during the years of study available in the first data release had some small differences due to its earlier phase of development. At the time, calls data were temporarily not collected, and the original “overall phone usage” measure was being used. In case of any onboarding effects, we excluded the first day of each participant’s time series. As a consequence, each participant’s time series started at midnight.

### Behapp data preprocessing

#### Phone usage-related channels

From the raw timestamps, the app and call-related activity channels were processed into hourly bins, showing the percentage of each hour a person spent carrying out each activity. Within each hour, these percentages were then binned to reflect “no activity” (0% of an hour spent carrying out the activity) or “activity” (>0% of an hour spent carrying out the activity). We used these bins due to the zero-inflated nature of the activity channels, as in Leaning et al. [25]. As the phone usage derived from screen status was developed later, for this study we processed screen status into hourly bins where each hour contained a list of all of the statuses the screen had had in the hour (including scheduled checks). If the “screen on and unlocked” status occurred in an hour we counted this as “Activity”, otherwise “No activity” was registered. Additionally, if “screen on and unlocked” was the last status in an hour and had no following “screen off” status in that hour, “Activity” was registered for the following hour under the assumption that the screen remained on and unlocked. For consistency with iOS users, we did not make use of the “screen on but locked” status recorded for Android users. We verified that this new overall phone usage measure had high agreement with the original phone usage measure present in Android users, with the two measures agreeing for >99% of hours.

To identify missing data, data from recurrent sampling-based sensors (including GPS and WiFi) was used, as the expected sampling frequency of these sensors was greater than once per hour. We could therefore differentiate between hours that had no recorded data due to missingness (NaN/NA), or due to an absence of activity (0% activity). Missing phone usage-related activity data were imputed according to the method in Leaning et al. [32], using Poisson Point Process Models (PPPMs). In this method, personalised PPPMs were fitted to each participant’s available data for each channel. We included a single covariate in these models, the hour of the day (encoded using one-hot encoding). The trained models could then be used to simulate data, with the simulations used to impute the missing values. The corresponding “hour of the day” values were provided to the PPPMs to ensure the correct values were used during simulation. As data can only be present in the other phone usage channels if there was overall phone usage, we constrained the imputation to only allow activity if the imputed overall phone usage value showed activity. We did not carry out imputation on the SMARD time series as a very small amount of phone usage was missing (2%).

#### Location-related channels

The GPS data were clustered to identify locations where participants were stationary (referred to as “staypoints”) [35]. The DBSCAN algorithm was used for clustering, with staypoints being defined as locations within a 150m radius where a participant remained for at least 30 minutes. Periods where a staypoint was not identified were then classified as periods when a participant was travelling. The location that was most visited between midnight and 6am was classified as a participant’s home location. The GPS data were then split into three main channels: home stay, stationary (excluding home) and travelling. Like the phone usage-related channels, these location channels also reflected the percentage of each hour during which the person was at each location (or travelling). The travelling channel was binned to show whether a person had travelled or not each hour (0% travelling; >0% travelling) as it was also zero-inflated, however the home stay and stationary channels were zero-one-inflated, and therefore split into three options (0%; >0 to <100%; 100%).

Two additional channels reflecting the different staypoints visited were also included in the MENTALPRECISION analysis. These channels were not included in the SMARD analysis due to the limited number of staypoints visited during their study participation, presumably due to the covid restrictions in place at the time. The original “staypoints visited” channel showed the staypoint/s that were visited each hour, with it being possible to visit a maximum of three staypoints each hour due to the staypoint parameters (i.e. starting the hour in a staypoint that was already being visited in the preceding hour, changing to another staypoint, and then going to another staypoint that continued into the following hour). The original encoding of the staypoints reflected the order in which they were visited throughout the study, which is not necessarily meaningful. We therefore re-encoded the staypoints to reflect the percentage of time participants spent at each of their staypoints, with the first staypoint being the staypoint where they spent the most time (which was usually “home”). Since participants could have a large number of staypoints, we binned the fourth+ staypoints all into one bin to reflect less commonly visited staypoints. A zeroth staypoint indicated that no staypoint was visited for the hour. We then split the original “staypoints visited” channel into two channels, reflecting the first staypoint visited each hour and the second staypoint visited each hour. A third staypoint per hour was visited so infrequently (for 0.07% of timepoints) that we did not include a column to reflect third staypoints.

Missing GPS data were identified separately to the remaining channels, using only the expected sampling frequency (>1/hour) of the GPS data. If GPS data were missing for an hourly bin, all of the GPS channels were marked as “NA” for that hour. We did not impute the missing GPS values as we did not deem our imputation method suitable for imputing GPS channels, as these behaviours generally occur more continuously than the phone usage-related behaviours. The “depmixS4” [36] package used in our analysis is able to handle missingness by omitting these values from the log-likelihood calculation, so this did not cause any issues in model training.

### Hidden Markov Model

The processed Behapp channels were used to train HMMs using the “depmixS4” [36] package in R. HMMs learn “hidden states” to represent the data, with each hidden state having corresponding probabilities of emitting the observed behaviours (emission probabilities), that are learnt during model training and can be used to interpret which observed behaviours each hidden state corresponds to. The observed behaviours were modelled using a multinomial distribution with an identity link function (equivalent to a binomial distribution for channels with binary values e.g. phone usage). At each timepoint, a participant occupies one hidden state, and the probability of transitioning between hidden states is learnt. DepmixS4 maximises the expected joint log-likelihood of the model parameters using the expectation-maximization algorithm. As in our prior work [25], we included dummy covariates encoding hour of the day (i.e. using one-hot encoding) on the transition probability formula, so that the probability of transitioning between states could depend on the time of day. We encoded the hour of the day as this is a cyclical covariate (ranging from 0 (midnight) to 23).

For both parts 1 and 2 of this study, a range of hidden state numbers were considered. After selecting the final HMM for each part, we generated the hidden state sequences corresponding to each participant’s time series using the Viterbi algorithm. The hidden state sequence is equal in length to the observed time series, meaning that it is possible to “transition” to the same hidden state between timepoints (i.e. if the participant stays in the same hidden state). We report the associated emission probabilities from the chosen HMMs, the hourly transition probabilities and the probability of starting in each hidden state.

### Part 1: non-clinical factors investigated using MENTALPRECISION

#### Smartphone usage and lifestyle questionnaire

To be able to understand additional factors that may influence an individual’s time series, we developed a questionnaire to record typical smartphone usage and lifestyle factors that could affect a participant’s digital behavioural patterns. Participants were interviewed before Behapp participation and authors LS, IW and MB recorded their responses.

The full questionnaire, containing 51 items (21 closed answer items, 30 open answer items), is provided in Supplementary Material 1. Many of the open answer items provide further detail on “Yes” responses to the closed-format questions, and so do not need to be answered if a “No” response was given. The SULQ had three parts, pertaining to phone usage, technology and lifestyle behaviours. For example, we asked participants questions about whether they used their phones for work (and if so, when), if they had an additional phone they used, if they used other technologies, what kind of area they lived in (e.g. city vs rural). To follow up, participants were asked two additional SULQ questions at the end of Behapp participation, “During the monitoring period, did anything out of the ordinary occur that was not part of your usual routine? (e.g. weekend trip, exam week, significant events like a job interview, significant personal news)” and “If yes, specify what and when”, in order to gauge whether their digital phenotyping time series may feature behaviours outside of their normal routine.

As this was the first usage of our questionnaire, we were not certain which items would turn out to be informative for understanding the participants’ behavioural patterns. For example, we asked participants if they were dependent on their smartphone, however many participants (n= 58/73) responded with “yes” to this question and so we did not use this item in our statistical analysis. For binary response items, we therefore went through each item to see which had a sufficiently balanced split in response to be investigated, given our modest sample size. The items with a sufficient balance for our analysis pertained to whether or not participants use another phone that is not their personal smartphone, whether they are addicted to some extent to their smartphone, whether they use their personal smartphone for work, whether they use a computer, tablet or console for gaming (under the assumption that during these periods there would be no phone activity) and whether they use a computer or tablet for communication. We also asked participants to provide an open-ended explanation of the area in which they lived. The majority of responses referred in some way to living in a village, suburb or a city (for example “small village”, “city”), so these descriptions were manually grouped into three classes: “village”, “suburbs”, “city”. We also predicted participant group and operating system (however these were recorded separately to the SULQ).

#### Questionnaire items from the Healthy Brain Study

Due to the overall study design and aim of MENTALPRECISION, we received access to a subset of the clinical questionnaires from the HBS [26]. From this subset we focused on questionnaires that may be relevant for our digital phenotypes, selecting the UCLA loneliness scale [37], Anxiety Sensitivity Index (ASI) [38] (trait anxiety) and Self-Report Inventory of Depressive Symptomatology (IDS-SR) [39]. The trait anxiety questionnaire was collected at one timepoint during the HBS, and the loneliness and depression questionnaires were collected at 3 timepoints across the HBS (though there was no overlap between these timepoints and the Behapp data collection). We therefore used the sum score from the trait anxiety questionnaire and the mean sum score from the loneliness and depression questionnaires in our prediction models.

#### HMM training

The HMM for part 1 was trained on the following phone usage-related channels: social media app, communication app, overall phone usage, incoming/outgoing calls, and on the following location-related channels: travelling, stationary, home stay, and first/second staypoints visited in an hour. An example time series is provided in Figure 3.

**Figure 3:**
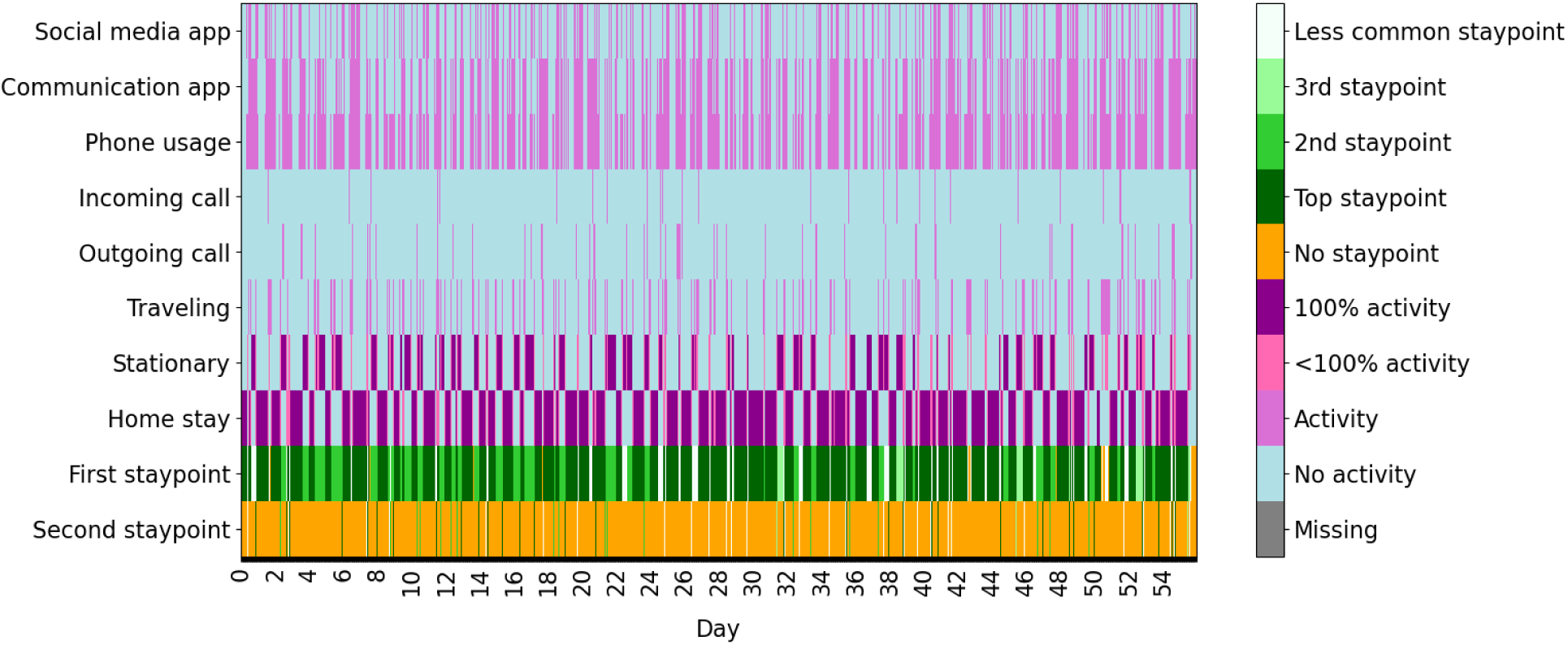
An example of the Behapp time series from a MENTALPRECISION participant, used in HMM training. Note that the “Activity” bin applies to the apps, phone usage, calls and travelling channels, whereas the “<100% activity” and “100% activity” bins apply to the stationary and home stay channels.

To select the number of hidden states used by the MENTALPRECISION HMM, we trained models with 2 to 7 hidden states and compared the Bayesian Information Criterion (BIC) between these models. We also considered the composition of the hidden states themselves by looking at their emission probabilities, to see whether there were redundancies in the hidden states. After selecting the number of hidden states we then considered the inclusion of individual transition coefficients (i.e. transition coefficients for each participant), so that the probability of transitioning between hidden states could be tailored to each participant. All other HMM parameters were shared between participants (i.e. group-based parameters). We used the parameters from the selected model in the updated model including individual transition coefficients. We initially attempted training the updated model with these parameters fixed, i.e. unable to be changed during training, so that only the individual transition coefficients were learnt. However the model failed to converge, so we therefore allowed all of the HMM parameters to be updated during model training. Finally, we compared the BIC between the HMMs with and without individual transition coefficients to select our final model.

#### HMM feature selection

We used the individual transition coefficients learnt by the HMM as features of interest in our prediction models. We converted these coefficients into probabilities so that we had an interpretable value for each possible transition and calculated the mean transition probability for each participant across the day. This then meant that for a HMM with *n* states, for each participant we had *n x n* mean transition probabilities. Another motivation for calculating probabilities from the coefficients was that the coefficients are determined by the HMM relative to a reference, which in our case is the first hour and first participant. If we used the transition coefficients themselves we would then not have suitable values for the participant treated as the reference due to the contribution from the reference hour.

After hidden state sequence generation, we calculated a digital phenotyping feature, the total dwell time, as in [25] and [32]. The total dwell times were the percentage of each participant’s time series that they spent in each hidden state (so therefore the number of total dwell times per participant was equal to the selected number of hidden states). We calculated the total dwell times over each participant’s entire time series as in this study we are investigating relationships between the digital phenotypes and measures that were made at single timepoints, rather than repeated measures that would warrant the dwell times to be calculated for smaller sections of their time series.

#### Visualisation

We then visualised the generated hidden state sequences, and carried out inspections of these hidden state sequences alongside the notes from the follow-up SULQ question. We present informative visualisations that show behavioural differences based on factors such as reported illness and holidays.

#### Regression analysis

We then investigated the relationship between the digital phenotyping features and SULQ survey and HBS outcomes. We ran two logistic regression models for each outcome: one where each of the total dwell times were included as predictors, and one where the mean transition probabilities were included as predictors (SULQ outcomes: whether or not participants use another phone that is not their personal smartphone, whether they are addicted to some extent to their smartphone, whether they use their personal smartphone for work, whether they use a computer, tablet or console for gaming and whether they use a computer or tablet for communication, area in which they lived; group, operating system; HBS outcomes: loneliness, trait anxiety, depression). As a number of the mean transition probabilities were small across all participants (close to 0%), we excluded these as they are not behaviourally meaningful. Given that there was high multicollinearity among the remaining mean transition probabilities, we carried out further selection of mean transition probabilities for use as predictors, as detailed in Supplementary Materials 2.

We included age as a nuisance covariate in all regression models. We carried out multiple comparison correction within each prediction goal, correcting for n-1 tests (where n is the number of hidden states) in models using the dwell times as predictors, and 14 tests in models using the transition probabilities as predictors.

### Part 2: relapse case study in SMARD

#### Relapse information

For the SMARD participant, we identified months containing relapse using the information recorded during follow-up, based on the clinical interviews (SCID). We ensured that these months were not included in HMM training and used the specific information regarding onset and recovery dates in our visualisations.

#### HMM training

As the majority of study participation took place during the covid era, we decided to exclude the portion of their time series that took place before lockdown measures were implemented in the Netherlands (23rd March 2020) [23]. We used the following three months to train the HMM, which importantly did not include a relapse and so were treated as a baseline/remission period. We did not include the encoded staypoints channels as the participant had a very small number of staypoints visited during their study participation (presumably due to the covid restrictions in place throughout their study participation). The model was therefore trained on the home stay, stationary, travelling, overall phone usage, communication app and social media app channels (see Figure 4). We considered 2-6 hidden states and selected the final number of hidden states based on the BIC of these HMMs. We included hour in the transition formula as in part 1.

**Figure 4:**
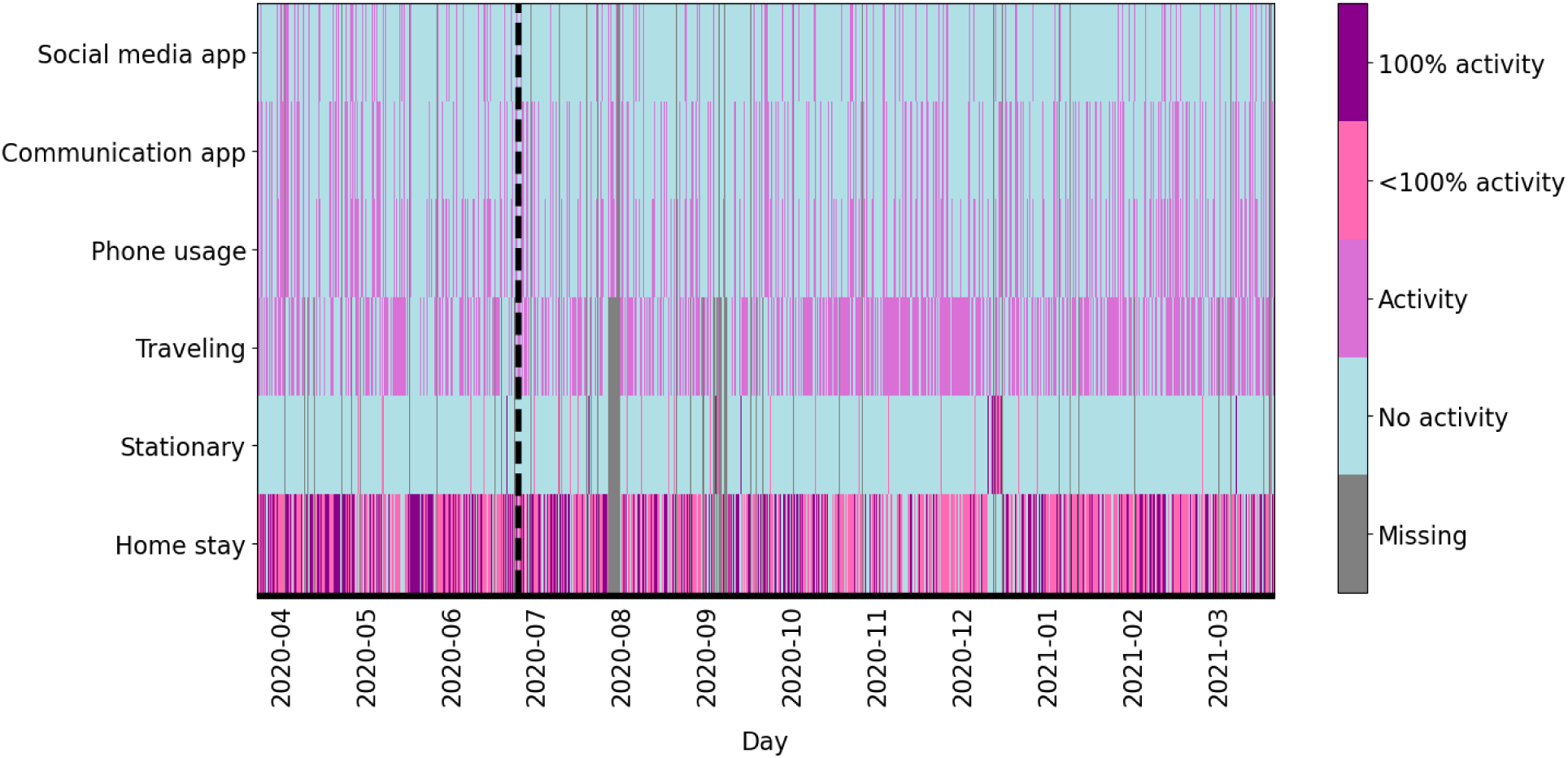
The Behapp time series from the SMARD participant. The first three months were used in HMM training, indicated by the dashed line.

#### Likelihood dynamics and behaviour

The time series was then split into monthly sections and the forward-backward algorithm run on each month, so that the likelihood of each month given the trained HMM could be extracted. Note that with this method the term “likelihood” does not refer directly to the likelihood of a relapse, rather the likelihood of a window of time given the participant’s baseline HMM. To avoid issues that occur when attempting to run the algorithm on months that do not contain certain values in their channels, the first month of the training series (specifically, the last week of March and entirety of April) was appended to each month to ensure that all values were present, so that the depmixS4 model object could be consistently created and the forward-backward algorithm successfully run. This then meant that whilst each month’s likelihood did not reflect purely its own data, every monthly likelihood was consistently impacted by the same portion of training data. The monthly likelihoods could then be compared to identify periods of lower likelihood. To demonstrate behavioural differences occurring over their time series, the Viterbi algorithm was run for the entire (training and withheld) time series to generate the corresponding hidden state sequence.

## Results

### Part 1

#### Sample statistics

At the time of analysis, 85 participants had been recruited for MENTALPRECISION. Two participants were excluded from the study as they did not meet our inclusion criteria. Two participants dropped out of the study and eight participants did not meet our data availability threshold. A histogram of data availability can be seen in Supplementary Materials 2 (Figure S1). Seventy-three participants met our criteria to be included in the analysis, of whom 32 participants had originally been recruited from the Healthy Brain Study cohort. The included participants had a mean age of 40 years (standard deviation = 15).

The majority of participants were HCs (n=57), with 16 cross-diagnostic participants. The sample included 36 males and 36 females (with one response not reported). Forty-one participants used an Android smartphone and 32 used an iOS smartphone. Forty-seven participants were employed, 9 were currently studying, 6 were currently studying and working, 5 were retired, 2 were unemployed and 4 reported other work-related situations. The sample had a mean number of years of education of 19 (standard deviation = 3), with 30 participants having attained a university education, 20 having attained secondary vocational education (“MBO” in Dutch), 20 having attained higher professional education (HBO), and 3 individuals each having attained pre-university education (VWO/gymnasium/atheneum), lower vocational education (LBO) and pre-vocational secondary education (VMBO).

#### Selected Hidden Markov Model

As the number of hidden states increased, the BIC decreased. However, when inspecting the six state model, it was seen that two of the states had very similar emission probabilities, and so we instead selected the five state model as all of its hidden states were notably distinct. When comparing the BIC of the five state models with and without individual transition coefficients, it was seen that the BIC favoured the model that included individual transition coefficients and so we selected this HMM for the remainder of the analysis. Whilst the emission probability parameters could be updated during refitting of the model with individual transition coefficients, these parameters did not change drastically. The emission probabilities from the selected HMM are shown in Figure 5. We labelled these states as follows: state 1 corresponds to a “socially active mixed locations” state, state 2 corresponds to “socially active home time”, state 3 corresponds to “less socially active stationary location”, state 4 corresponds to “less socially active travelling and/or home” and state 5 to “socially inactive home time”. By saying “less socially active”, we are referring to the probability of social activity being lower for states 3 and 4 than states 1 and 2. The transition probabilities, calculated with respect to the reference participant, are shown in Figure 6. For all states except state 1, the probability of transitioning within the same state was higher throughout the day. The consecutive starting probabilities for states 1 to 5 were 0.014, 0.234, 0.069, 0.026 and 0.657, i.e. it was most probable that a participant would begin their time series in state 5.

**Figure 5:**
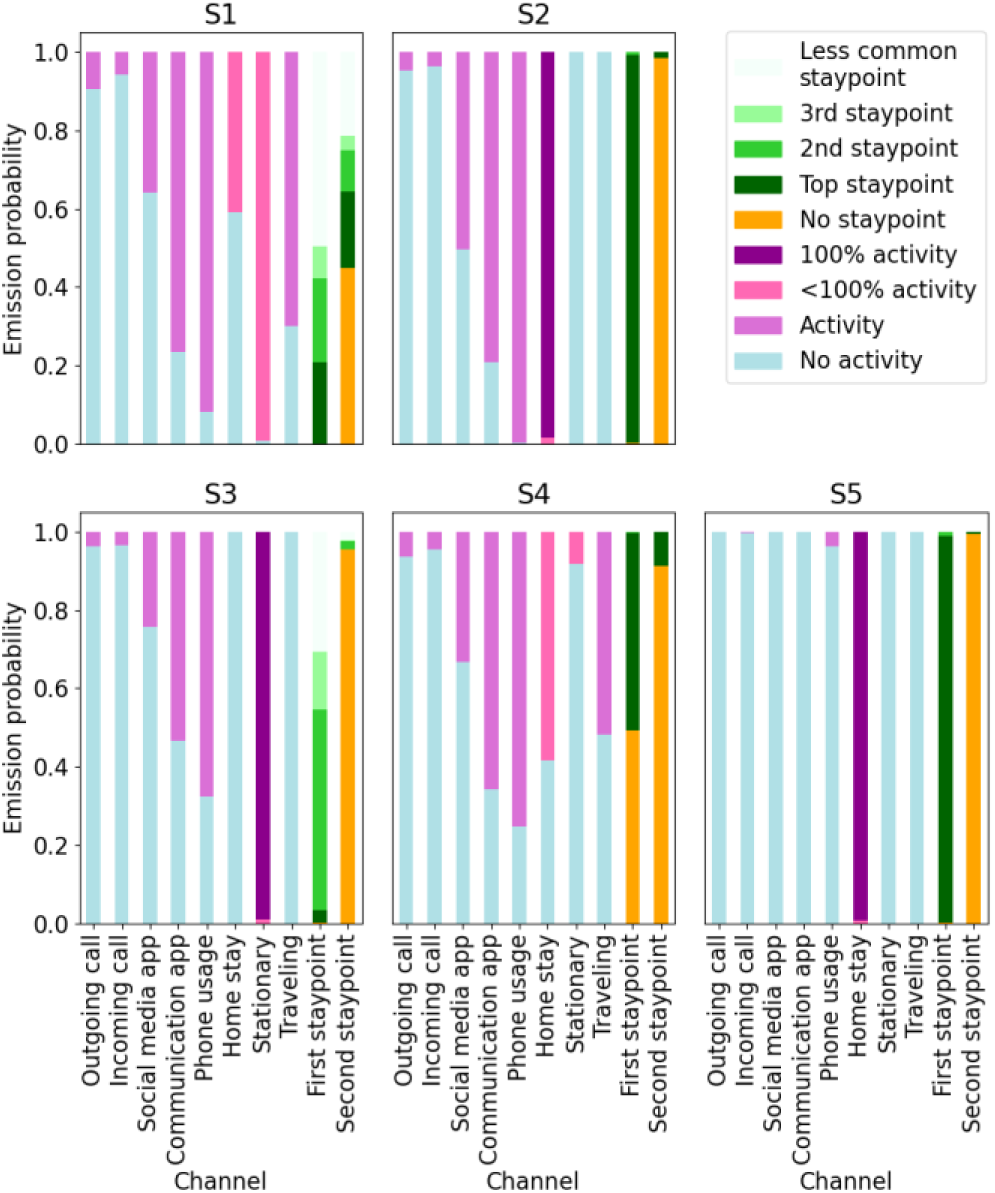
The emission probabilities from the selected 5-state HMM. S: state. S1: “socially active mixed locations”, S2: “socially active home time”, S3: “less socially active stationary location”, S4: “less socially active travelling and/or home”, S5: “socially inactive home time”.

**Figure 6:**
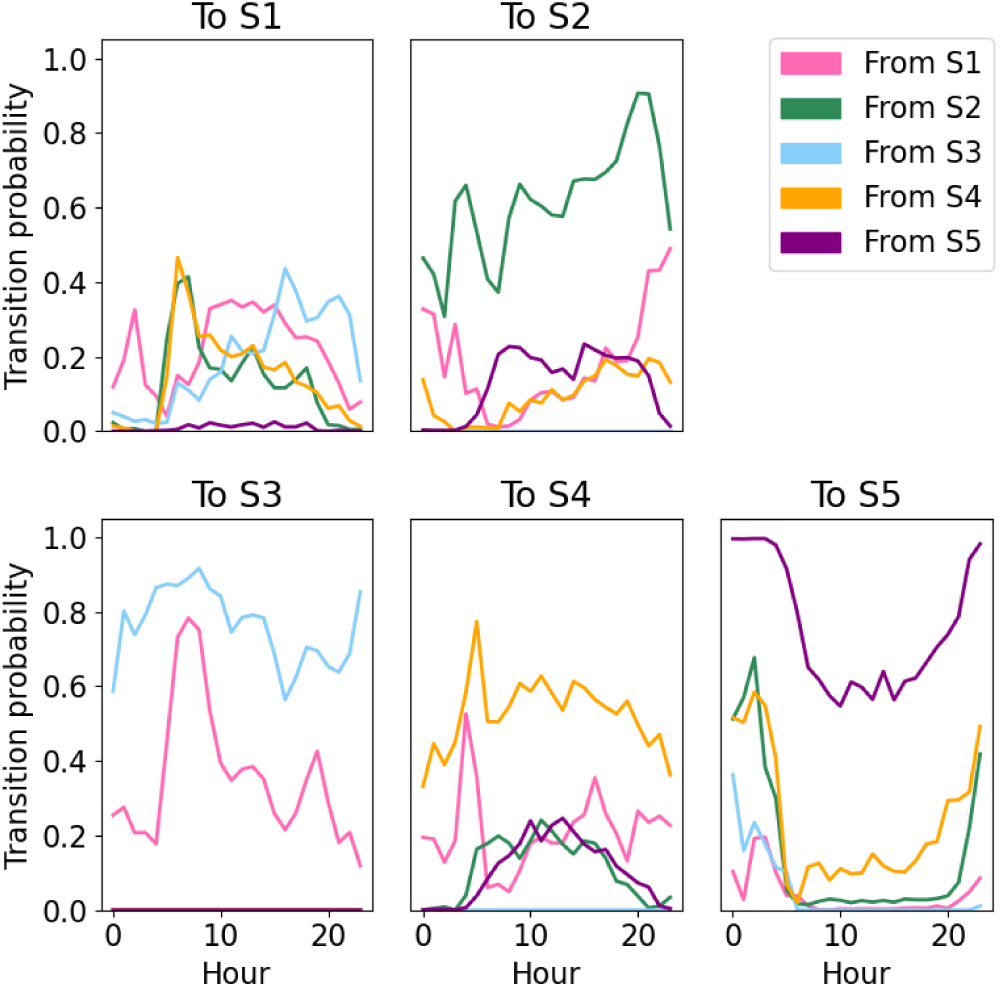
The transition probabilities from the selected 5-state HMM, given the reference participant. Each subplot shows the transitions to a given state from all of the possible states. S: state.

#### Visualisations using SULQ

In the post-study interview, several participants reported periods of illness or holidays they had taken. These periods are evident in the visualisations of their hidden states. The participant in Figure 7, reported that they were physically unwell during the last few weeks of study participation, and the switch from well to unwell can be clearly seen in their hidden state sequence. Before the onset of their illness, they had a regular daily behavioural pattern that involved social activity and leaving the home in the morning, a period away from home during the day, generally matching their self-reported work hours (described as being at their work place on Monday-Thursday from 9.00-14.00 and Friday from 9.00-13.00), and a socially active return home. This then dramatically switched to much less periods spent away from home once their illness had begun. The participant in Figure 8 reported going on holiday during “the end of July” and this holiday period is clearly evident in the middle of their time series, with the majority of time spent in the stationary and mixed location states. Figure 9 shows the pattern of a person who works from home on Monday-Thursday, showing much more time spent in home-related states than people working away from home (e.g. Figure 7 before illness onset).

**Figure 7:**
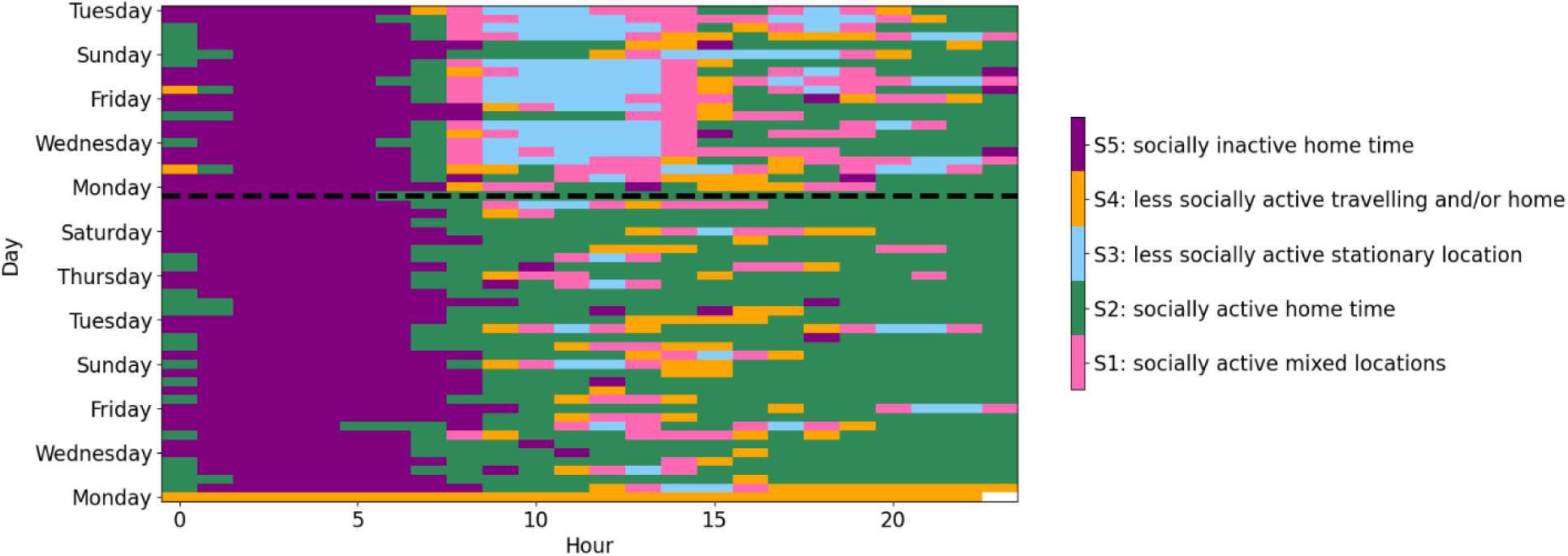
A participant who was unwell during the last few weeks of study participation. Approximate illness onset is indicated by the dashed black line, plotted after three weeks of study participation.

**Figure 8:**
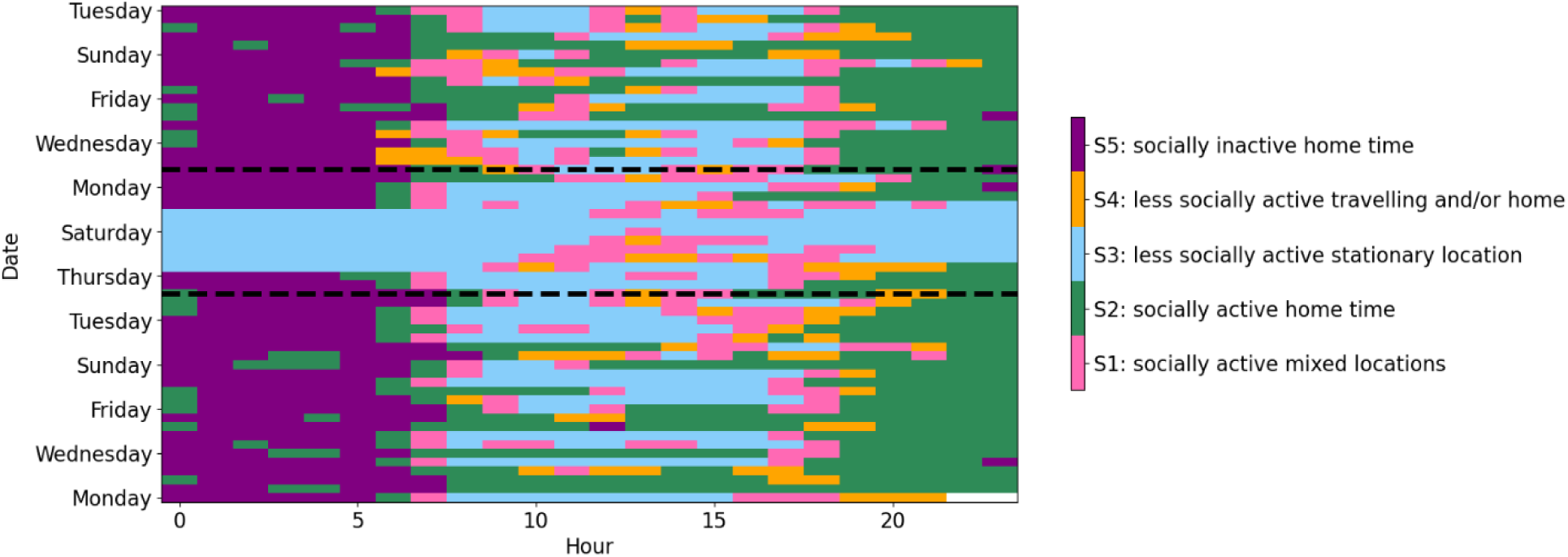
A participant who went on holiday during the middle of study participation (“end of July”). The dashed lines indicate the midpoint of July and the end of July. It can be seen that the majority of this time was spent in state 3 (blue).

**Figure 9:**
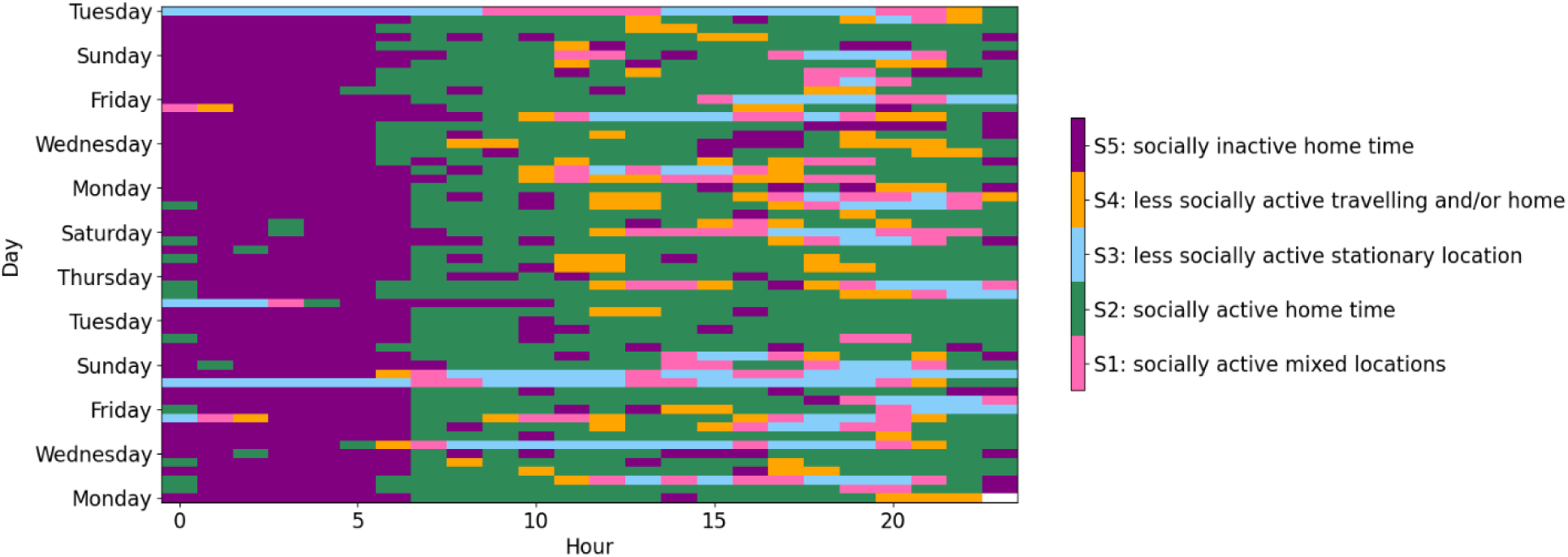
A participant who works from home. During the day this participant spends a lot of time in state 2 (green).

#### Regression analysis

We carried out several regression analyses to investigate possible relationships between the digital phenotypes and the SULQ measures, as well as clinical outcomes from the HBS. Significant relationships between various total dwell time predictors and the “other phone”, “operating system” and “smartphone addiction” outcomes are shown in Figure 10.

**Figure 10:**
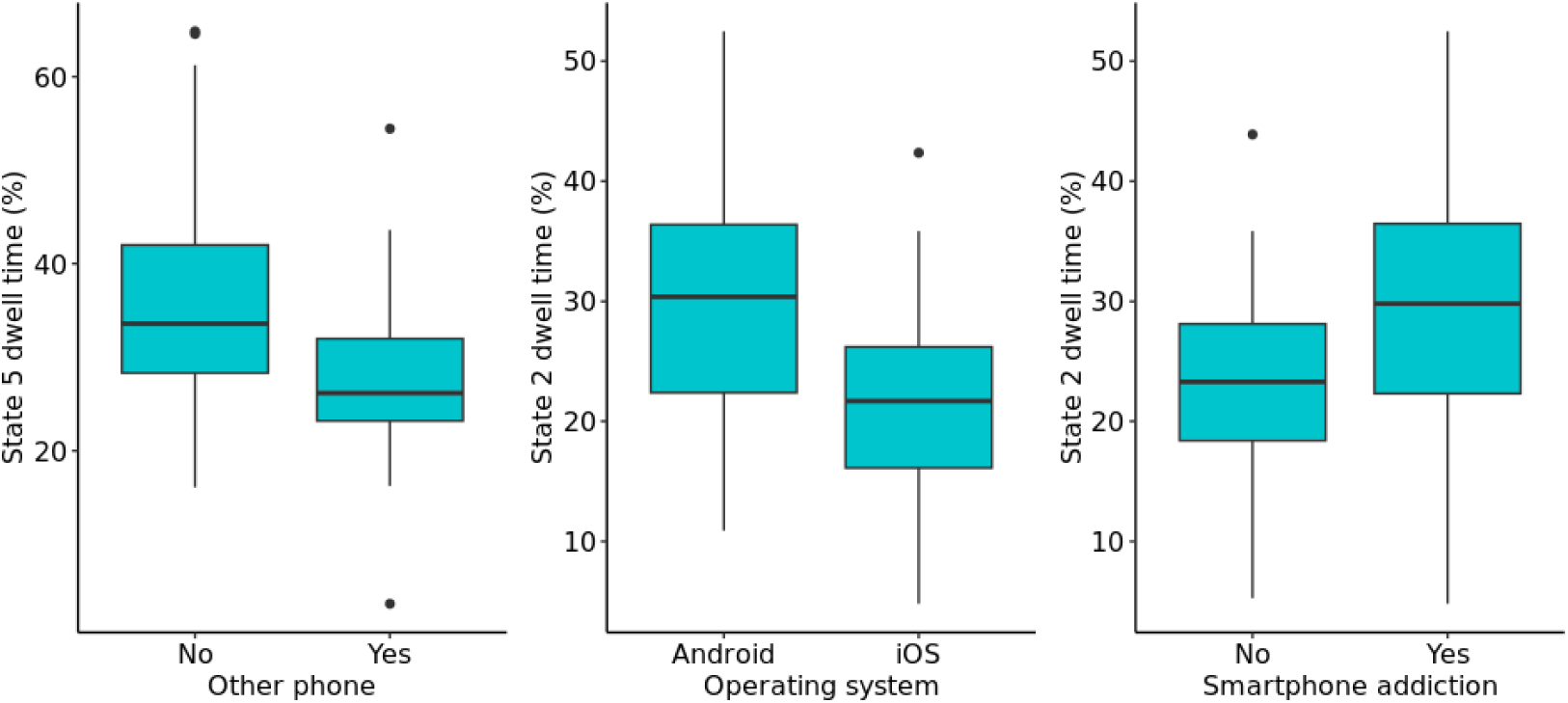
Box plots of the significant dwell time predictors for the “other phone”, “operating system” and “smartphone addiction” outcomes.

A logistic regression model was run to investigate if there were significant differences in the total dwell times of participants who had one phone versus participants who also use another phone (in most cases for work purposes), which could be driven by differences in phone usage. Results are presented in Table 1, with FDR correction of *P* values carried out considering 4 tests and a significance threshold of <.05. There was a significant difference in the state 5 (“socially inactive home time”) total dwell time (FDR-corrected *P*=0.03, odds ratio 0.9196, 95% CI 0.8583-0.9808), with participants with an additional phone spending less time in this state. This could reflect differences in work commitments for participants with multiple phones, impacting their social activity levels outside of work.

**Table 1:** Predicting whether participants have another phone using the total dwell times. Number of “no” responses=48, “yes” responses=25. “No” is the reference class.

| Dwell time predictor | Coefficient | Standard error | Odds ratio | CImin | CImax | z value | P value | FDR-corrected P value |
| --- | --- | --- | --- | --- | --- | --- | --- | --- |
| State 1 | 0.1363 | 0.0744 | 1.1460 | 1.0001 | 1.2919 | 1.8307 | 0.07 | 0.27 |
| State 2 | 0.0176 | 0.0242 | 1.0177 | 0.9703 | 1.0651 | 0.7261 | 0.47 | 1.00 |
| State 3 | -0.0467 | 0.0352 | 0.9544 | 0.8853 | 1.0235 | -1.3244 | 0.19 | 0.74 |
| State 4 | 0.0122 | 0.0226 | 1.0123 | 0.9680 | 1.0565 | 0.5396 | 0.59 | 1.00 |
| State 5 | -0.0839 | 0.0313 | 0.9196 | 0.8583 | 0.9808 | -2.6826 | 0.007 | 0.03 |

We then considered possible differences between participants with Android and iOS smartphones (Table 2). We saw a significant difference in the state 2 dwell times (“socially active home time”) with participants with iOS smartphones spending significantly less time in this state (FDR-corrected *P*=0.04, odds ratio 0.9330, 95% CI 0.8804-0.9857), which could reflect behavioural differences between users of different operating systems, or differences in the operating systems themselves.

**Table 2:** Predicting operating system using total dwell times. Number of participants using an Android=41, number of participants using an iOS=32. “Android” is the reference class.

| Dwell time predictor | Coefficient | Standard error | Odds ratio | CImin | CImax | z value | P value | FDR-corrected P value |
| --- | --- | --- | --- | --- | --- | --- | --- | --- |
| State 1 | -0.1334 | 0.0749 | 0.8751 | 0.7284 | 1.0219 | -1.7817 | 0.07 | 0.30 |
| State 2 | -0.0693 | 0.0269 | 0.9330 | 0.8804 | 0.9857 | -2.5805 | 0.00987 | 0.04 |
| State 3 | 0.0350 | 0.0331 | 1.0356 | 0.9708 | 1.1004 | 1.0575 | 0.29 | 1.00 |
| State 4 | 0.0506 | 0.0348 | 1.0519 | 0.9836 | 1.1201 | 1.4513 | 0.15 | 0.59 |
| State 5 | 0.0630 | 0.0327 | 1.0650 | 1.0009 | 1.1291 | 1.9250 | 0.05 | 0.22 |

We then investigated self-reported smartphone addiction (Table 3). Participants reporting a smartphone addiction spent significantly more time in state 2 (“socially active home time”) (FDR-corrected P=0.009, odds ratio 1.0787, 95% CI 1.0301-1.1272) than participants reporting no smartphone addiction. This could reflect their increased time spent on their phone, when they are at home not occupied by other commitments, as a result of their behavioural addiction.

**Table 3:** Predicting smartphone addiction from total dwell times. Number of “yes” responses=40, “no” responses=33. “No” is the reference class.

| Dwell time predictor | Coefficient | Standard error | Odds ratio | CImin | CImax | z value | P value | FDR-corrected P value |
| --- | --- | --- | --- | --- | --- | --- | --- | --- |
| State 1 | -0.1343 | 0.0712 | 0.8744 | 0.7348 | 1.0139 | -1.8858 | 0.06 | 0.24 |
| State 2 | 0.0757 | 0.0248 | 1.0787 | 1.0301 | 1.1272 | 3.0580 | 0.002 | 0.009 |
| State 3 | 0.0348 | 0.0312 | 1.0354 | 0.9743 | 1.0965 | 1.1166 | 0.26 | 1.00 |
| State 4 | 0.0093 | 0.0217 | 1.0093 | 0.9669 | 1.0518 | 0.4278 | 0.67 | 1.00 |
| State 5 | -0.0253 | 0.0234 | 0.9750 | 0.9292 | 1.0208 | -1.0831 | 0.28 | 1.00 |

We investigated possible differences in the digital phenotypes between participants living in a city (the reference category) and participants living in a village or suburbs using multinomial logistic regression, and no significant differences were observed. We also considered whether participants use their phone for work, whether participants use a computer, tablet or console for gaming, and whether they use a computer or tablet for communication, and no significant relationships were identified.

For participants from the HBS study, we also investigated clinical outcomes (mean loneliness score (ULS) (n=30), mean depression score (IDS) (n=30) and the sum trait anxiety score (ASI) (n=29)), and for all participants (n=73) we investigated their classification as being part of the healthy or cross-diagnostic patient cohorts. No significant relationships were identified for any of these outcomes when using the total dwell times for each state as predictors.

We then investigated the selected mean transition probabilities as predictors for each of these outcomes. Whilst several of these results were significant prior to multiple comparison correction, none of these results survived the multiple comparison correction.

### Part 2

#### Sample statistics

The selected participant had a total study duration of 548 days, participating from October 2019 until March 2021. They experienced one depression relapse, occurring about three quarters of the way through study participation (after approximately 14 months), reported as occurring from mid-November until the first week of January. Demographics are not presented for this participant to protect their anonymity.

#### Selected Hidden Markov Model

Based on the BIC of the candidate models (considering 2-6 hidden states), a 3-state HMM was selected. The hidden states of this model, interpreted from the emission probabilities (Figure 11), corresponded to a socially inactive (home and) travelling state, a socially inactive home state and a socially active home (and travelling) state. The participant started their time series in state 2. Their transition probabilities are shown in Figure 12.

**Figure 11:**
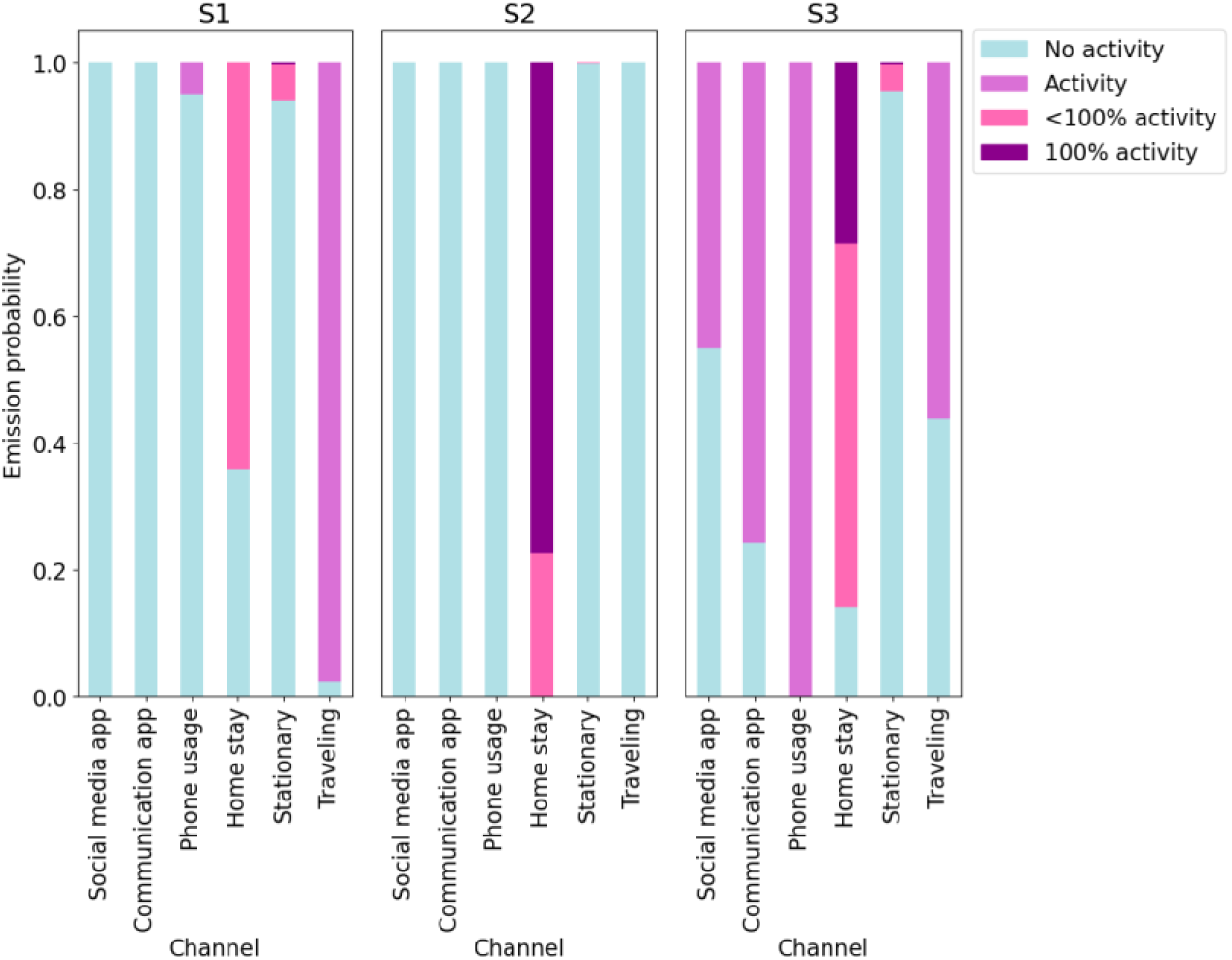
Emission probabilities for the SMARD case study. S1: “socially inactive (home and) travelling”, S2: “socially inactive home time”, S3: “socially active home (and travelling)”.

**Figure 12:**
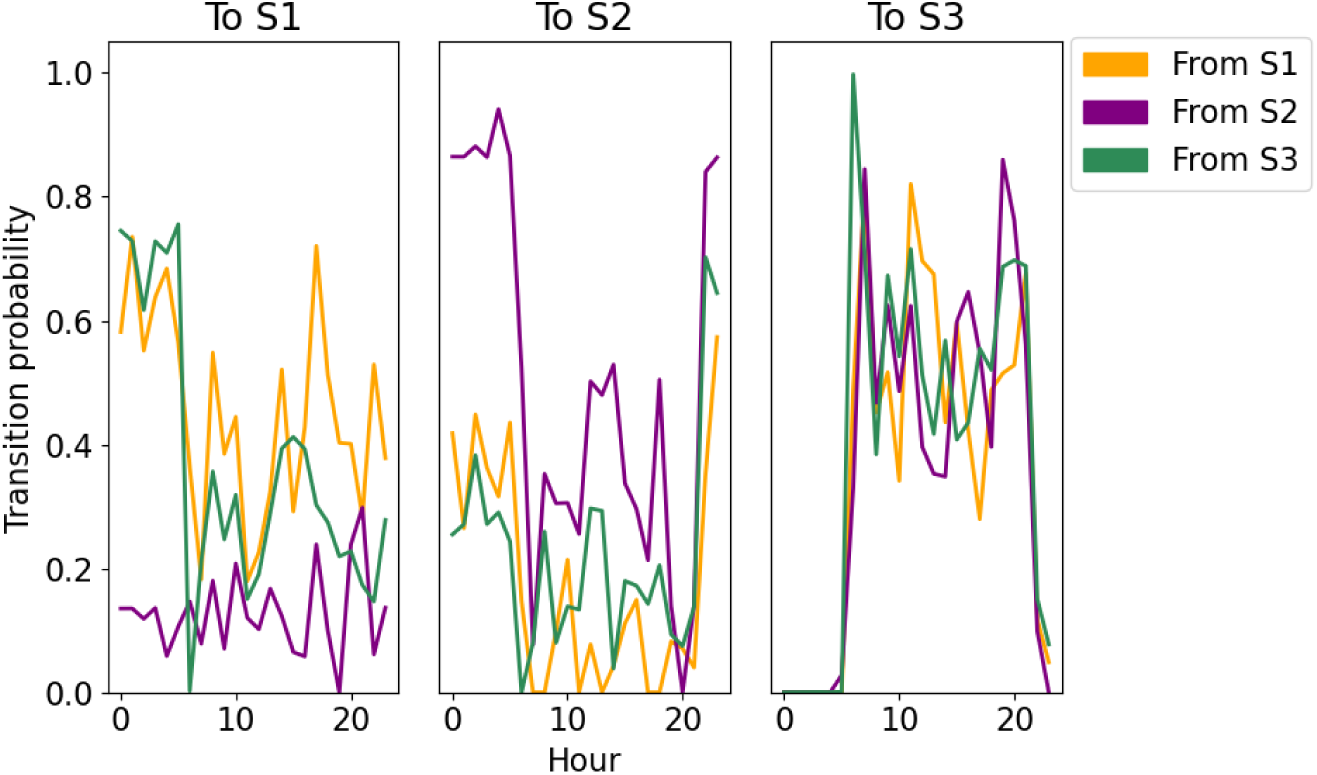
Transition probabilities for the SMARD case study.

#### Likelihood dynamics and behaviour

Figure 13 shows the month-by-month likelihoods of the time series. It can be seen that the likelihood decreased around October-November-December 2020, being consistently lower for these months, before starting to increase again in January 2021 and returning to what appears to be normal in February. This dip coincides with the participant’s relapse, which was reported as beginning in mid-November and lasting until the first week of January. This pattern is consistent with our hypothesis that depression relapse periods can be identified as periods with lower likelihood, given a HMM trained on a period of remission.

**Figure 13:**
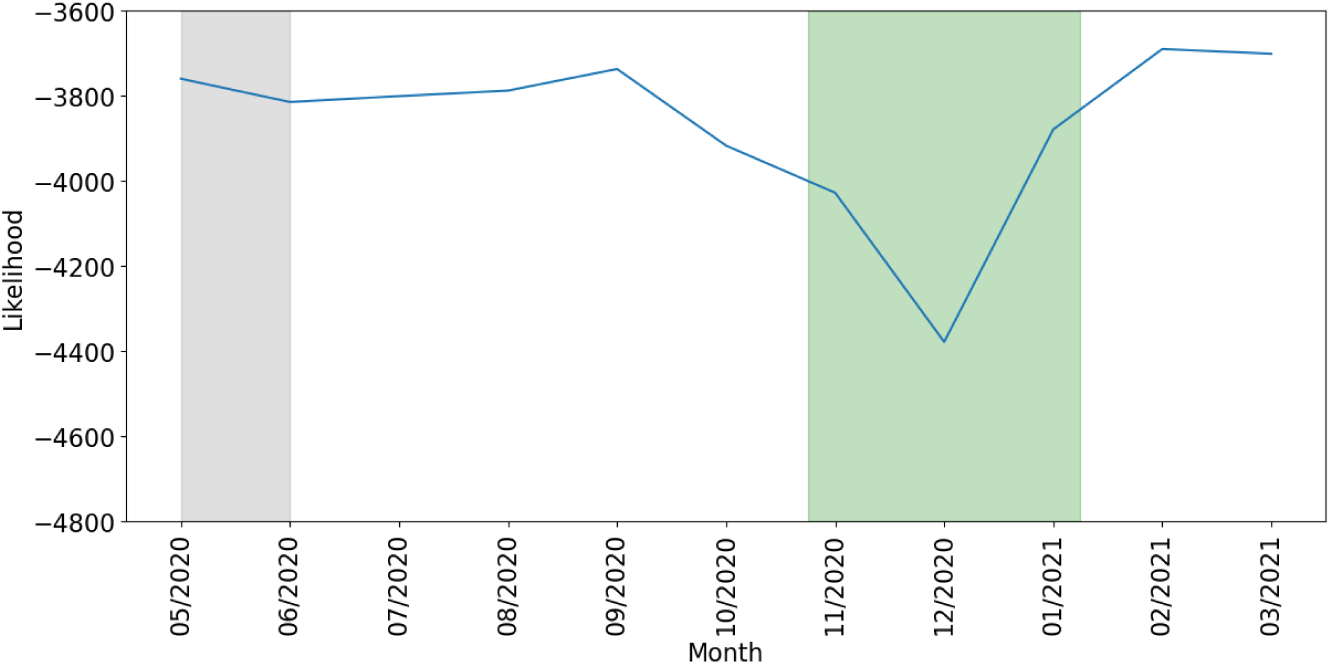
The month-by-month likelihood of the time series given the trained model. The grey shading indicates months that were used for training (with the first training month excluded as this was the supplemental month used to ensure the algorithm would run successfully), and the green shading indicates the approximate relapse period.

This participant’s hidden state sequence is displayed in Figure 14, where each row corresponds to one day. Behavioural changes preceding and during the relapse can be seen, notably that during the nighttime the participant was occupying the socially inactive (home and) travelling state more. This suggests that around this period the person had more disrupted nighttime periods than what is normal for them, spending more time outside the home.

**Figure 14:**
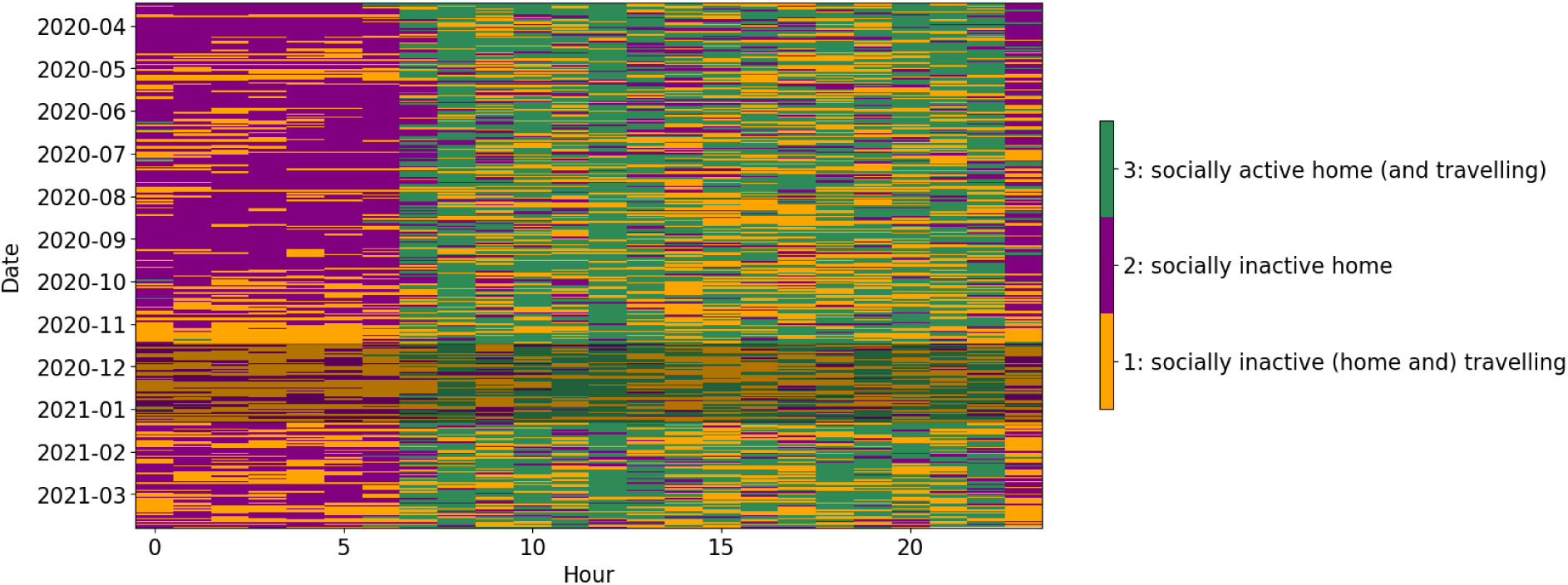
The hidden state sequence generated by the HMM trained on baseline data. The shaded area corresponds to the relapse period, occurring from mid-November until the first week of January.

## Discussion

### Principal results

In this study we investigated non-clinical factors that may be relevant for improving personalised digital phenotyping models, and considered methods to address the temporal nature of digital phenotyping. We considered these elements in one of the key clinical goals of digital phenotyping: depression relapse prediction.

We compared the MENTALPRECISION hidden state sequence visualisations to SULQ responses and saw clear behavioural changes around reported illness and holiday periods, showing how it is important to take into account other factors that may affect an individual’s behaviour before drawing clinical conclusions. Our logistic regression results showed several statistically significant differences between different self-reported lifestyle and phone usage behaviours, with the dwell time for the “socially inactive home time” state being significantly lower for participants who use another phone in addition to their personal smartphone. The dwell time for the “socially active home time” was significantly lower for iOS users than Android users, and significantly higher for participants reporting a smartphone addiction when compared to participants reporting no smartphone addiction. This suggests that digital phenotypes can be associated with non-clinical outcomes related to a person’s habits. Digital phenotyping researchers should therefore be mindful of the possibility of measuring non-clinical signal, and that there is a risk of attributing this to clinical factors. We recommend that future studies acquire information surrounding phone use habits so that non-clinical factors can be included in digital phenotyping models to reduce this risk.

Finally, we showed a proof-of-concept application to illustrate the use of digital phenotyping measures to make time-resolved predictions of relapse in recurrent depression. In this analysis, we noted a consistently lower likelihood score in the months containing a relapse, during which higher levels of nighttime activity were observed.

#### Methodological contributions

We provide contributions to improving personalisation in digital phenotyping through evaluating non-clinical factors and HMM modelling decisions. We evaluated non-clinical factors in part 1 of the study, where we investigated the relationship between SULQ measures and digital phenotypes, identifying various significant relationships and obvious behavioural differences coinciding with reported events occurring during study participation. This indicates that future studies should give care to ensuring that effects detected in digital phenotyping data correspond to clinically relevant effects rather than more fundamental changes in routine (e.g. physical illness, going on holiday). During SMARD data collection, the SULQ had not yet been created. However, the data collection for this participant did take place during covid, an obvious example of a potent non-clinical factor influencing behaviour (for example as seen in [40]). It was seen that a small number of hidden states were appropriate for this participant, possibly influenced by the covid rules in place at the time, as the learnt hidden states were not associated with a large probability of stationary time (Figure 11). This contrasted with the HMM in part 1 where different states reflected different probabilities of exhibiting stationary time. We also note that whilst the likelihood was lower for multiple months, it was at its lowest in December. Considering the original Behapp sequence (Figure 4), it can be seen that there was a prolonged period of away time during this month, possibly due to the seasonal holidays, which could be a possible example of the combination of clinical factors (depression relapse) and non-clinical factors (holidays).

In part 1 of the study, we incorporated a level of individualisation in the HMM by allowing for the state transition probabilities to depend not only on the hour of the day but also on participant ID. This improved the BIC of the HMM and also provided us with an additional HMM-derived phenotype, in the form of individual transition probabilities. In part 2 of the study, the HMM was entirely personalised to one participant, with comparisons made between baseline and relapse periods instead of between subjects.

Given the clinical goals of digital phenotyping related to symptom and relapse prediction, digital phenotyping models should aim to provide a time-resolved analysis of behaviour. This has proved challenging, as many studies do not collect high temporal resolution validation metrics and is further complicated by the timing challenge of recording data over periods when a relapse occurs. It is therefore difficult to associate changes in digital phenotyping time series with clinical endpoints. Studies often end up comparing summary measures of digital phenotypes to the clinical endpoints, or coming to somewhat of a compromise by calculating summary measures that reflect temporal properties [10], for example circadian rhythm. In part 1 of this study, we introduced a similar “compromise” measure by including individual transition probabilities in the HMM, to reflect the dynamical properties of the time series and then using these as a predictor in our statistical models. In part 2 of this study, we took further steps toward time-resolved analysis by evaluating the likelihood of the time series month-by-month, thereby identifying monthly changes in their time series. In this case study, this approach shows clear, albeit preliminary, potential of the ability of digital phenotypes to signal an episode of mental illness. However, we acknowledge that this should be replicated in additional data.

### Future directions

We have shown evidence for relationships between non-clinical factors and digital phenotypes in a predominantly “healthy” sample. These relationships can also be investigated in a clinical sample to investigate how clinical and non-clinical signals combine in digital phenotyping time series. Future studies should investigate how to appropriately include non-clinical factors in digital phenotyping models. For example, they could be included as covariates in the HMM (as the participant ID and hour were included in our HMMs), or as additional covariates in the statistical models when investigating relationships with clinical measures (as we did with the age nuisance covariate). These factors could also be incorporated into novel measures. For example, to determine the risk of relapse, the HMM-provided likelihood measure could be combined with SULQ measures to provide a composite score, or the SULQ measures could be used to vary a threshold for relapse detection. For example, if a participant reports a physical illness then the threshold for identifying concerning likelihood scores could be made more generous so that a possible relapse is less likely to be flagged. As this was the first use of the SULQ, its psychometric properties need to be validated, for example to ensure good test-retest reliability.

We have shown a proof-of-concept for using the HMM-derived likelihood of a time series to identify depression relapse. After validating the method in a larger sample, once newer SMARD data using the more recent Behapp version has been processed, future investigations should fine-tune analysis details, such as identifying how much of a decrease in likelihood (and for how long) is indicative of relevant behavioural changes (for relapse threshold setting), as well as a more optimal window construction for likelihood calculation, considering what length of prediction window (e.g. [41]) and time-resolved approach (such as rolling windows) is appropriate for optimal relapse detection.

Our proof-of-concept approach focused on a completely individualised model, in part due to the data availability issues we experienced. A model including both group and individual parameters (such as in the MENTALPRECISION part of this study) could also be investigated and compared to purely individual models, to determine whether this method may also allow relapse to be identified using a likelihood score.

#### Recommendations

We have found the SULQ to be a useful tool in interpreting and analysing digital phenotyping data. We designed the SULQ based on information that we felt was potentially important, but lacking, when we were interpreting behavioural patterns from previous studies. For example, sometimes we noticed behavioural changes in the data but had no idea what occurred at that time in the participant’s life. Additionally, it is plausible that many observed behaviours could have either a positive or a negative cause, for example if a person spent a long period at home, this could be because they were unwell, or because they were relaxing at home during a holiday. Without knowing any other context for the observed behaviour, we were cautious to attribute too much clinical significance to the behavioural patterns, and therefore developed and included this lifestyle and smartphone usage questionnaire in our study.

We encourage other researchers to collect lifestyle and smartphone use measures using an instrument such as the SULQ to assist in interpreting digital phenotyping time series and/or to improve their modelling approaches. We understand that different studies have different resource constraints, and therefore propose that if researchers prefer to streamline their data collection, that they focus on collecting the binary items, as it is more time consuming to record the open format questions. Any open format questions could then be selected depending on the research question. We collected the SULQ during interviews, however the questions could also be gathered using an active smartphone app, facilitating repeated measures.

Regarding the SULQ_area item (“What kind of area do you live in? (e.g. rural, city, suburbs)“), many participants provided easily labelled answers (such as “village” or “city”), although we did encounter some harder to label answers (e.g. “outskirts of a city”, which was labelled as “suburbs”). For our initial use of this survey, we wanted to allow participants flexibility in their answers, understanding that the area in which someone lives can be very different to someone else even if the same label would be appropriate. However, for future survey iterations a multiple choice question, allowing comments, could be included for easier labelling. Alternatively, classification could be done based on objective information such as postcodes. Whilst we did gather open format responses regarding work routines (including days worked from home), researchers could instead ask a multiple choice question such as “From where do you normally work?” with responses “A fixed location (e.g. office)”, “A non-fixed location”, “home”, or “other”, as we did not feel that the open format question allowed for sufficiently convenient investigation of a factor that may have a large impact on a person’s behaviour.

Alongside many of these binary/fixed response items, we collected unstructured answers to further elaborate on each participant’s routine. Whilst we expected these responses to be more challenging to use, they were in fact even more challenging than expected. There were large amounts of variation in how detailed each provided response was, with routines varying greatly between people. We expect that it will generally not be practical to extract data from these unstructured responses for use in statistical analysis, but they may help in the interpretation of individual behavioural patterns in more unstructured settings (e.g. the Digital Clinic [20]). A more structured question may also help to standardise the responses.

### Limitations

We showed through visualisations how periods such as holiday and illness provide overtly different behavioural patterns, but do not exclude these periods from our regression analyses, and so for some participants their digital phenotypes may not be reflections of their routine behaviours. It could be considered to exclude periods such as these from analysis, although this would require even more precise recording of these periods. It is also difficult to determine what would be someone’s “normal” behaviour, versus to-be-expected fluctuations in behaviour. For example, a holiday of one month could quite fairly be considered out-of-the-ordinary for most people, but would a night away similarly be considered out-of-the-ordinary? Efforts may therefore be better spent on further improving individualisation, such as in part 2. For goals such as relapse prediction, it would be expected that these out-of-the-ordinary periods would also be flagged as having a low likelihood score and may be potentially flagged for relapse. The nature of this period could then be resolved through a brief phone call, or to maintain automation, through a triggered EMA question (e.g. [29,42]).

A key aim of our study was to investigate possible lifestyle and phone usage factors that may impact digital phenotypes, however we focused on associations not causality. The SULQ factor may not necessarily cause the digital phenotype, as it could be that the measured factor has an underlying cause. Whilst we would like to draw attention to the possibility that supposed clinical differences could be due to (or related to) additional, non-clinical factors, by that same logic it could be that the differences we draw attention to are not due to the non-clinical factors we assessed after all.

Whilst we propose that collecting additional measures, such as those in the SULQ, will be important for the development of digital phenotyping, these measures are susceptible to the typical limitations of self-report, subjective measures. Within the categories we predicted, there could also be large between-person variation. For example, for participants with more than one phone it can greatly vary how much a person uses each phone. Participants may also have varied in their interpretation of questions regarding “dependence” and “addiction”, with some participants requiring these words to be defined during questionnaire completion. Additionally, to interpret the SULQ_area labels appropriately (“city”, “village”, “suburbs”), we would like to draw attention to the fact that the Netherlands has extensively developed infrastructure, with many smaller Dutch towns and villages having buses connecting them with more populated areas, or being within cycling distance (given the country’s small size) to towns and small cities with key services. Any behavioural differences may be more overt in countries with different infrastructures, for example in countries where rural areas are not reachable by public transport. Future work could also consider acquiring region-specific environmental data, using the GPS data collected by Behapp, in order to better understand regional variations in living environments [43].

The MENTALPRECISION study was able to include participants with iOS, not just Android, smartphones, due to advances in the Behapp application. However, we were unable to include app usage measures for iOS users. As phone usage was necessary for these channels to be active, we hope that this did not have a large effect on model learning and hidden state sequence generation, although this does highlight the difficulties in working with more than one operating system. We extracted transition probabilities from the HMM, however these had high multicollinearity and did not turn out to be significant predictors of our outcome measures. For more specific research questions, researchers may wish to focus on probabilities for specific transitions of interest.

In both of our HMMs, we have assumed that properties stay constant over time, for example the hourly probabilities of transitioning between hidden states. In reality, model parameters may change over time. Our likelihood method does not, however, depend on the stationarity of model properties as we capitalise on model fit to detect anomalous periods. On a related methodological note, we acknowledge that the differences in likelihood for the relapse period suggest that the trained HMM is a less suitable fit for this part of the time series, and so may not be an optimal indicator of the underlying hidden states for this period which are displayed in the visualisation. Nevertheless, we find the visualisation a useful demonstration of behavioural changes occurring around a relapse, which is difficult to achieve otherwise due to the dimensionality of the observed channels (see Figure 4). Whilst the likelihood may be a less interpretable feature than other HMM features that we have used, such as the total dwell time, the flexibility of this approach for handling unseen behaviours holds strong appeal. For a clearer idea of the underlying states occupied by a participant during these periods of identified behavioural change, the HMM could also be retrained for these periods and the new hidden states interpreted.

Note that we cannot make any causal inferences about the observed behavioural changes and relapse, leaving open questions: is the increased nighttime activity seen before the onset of the relapse an early warning signal of depression, or could it be a behavioural change that precipitated the onset of a relapse? Without further context, interpreting the increased nighttime activity is difficult. However, whilst there were varying covid restrictions during study participation (including lockdown measures over the relapse period), the visibly increased nighttime activity seems more likely to be related to depression than covid restrictions, especially as the lockdown continued well into 2021, when the month-by-month likelihood had increased again.

## Conclusion

Digital phenotyping is a developing approach to monitoring mental health symptomatology, but in focusing on clinical outcomes the field has often overlooked other factors that may impact digital phenotypes and be relevant for personalisation. Using a questionnaire designed to assist in understanding non-clinical factors impacting digital phenotyping, we observed how changes in behaviour could be seen in the digital phenotyping patterns, and identified significant differences between non-clinical measures. We also demonstrated a proof-of-concept for identifying depression relapse using a likelihood score provided by a personalised HMM. Improving personalisation in digital phenotyping through including personalised model parameters and sufficiently accounting for non-clinical factors may enable the improved detection of key clinical outcomes such as depression relapse. This could have important implications such as enabling patients and clinicians to determine when earlier treatment is needed.

## Supporting information

Supplementary Materials 1: the complete Smartphone Usage and Lifestyle Questionnaire.

Supplementary Materials 2: additional figures and further methodological information regarding multicollinearity of the mean transition probabilities.

## Acknowledgements

The MENTALPRECISION study was funded by the European Research Council (consolidator grant 101001118). The SMARD study was funded by the Hersenstichting (Dutch Brain Foundation) (HA2015.01.07).

## Data availability

The datasets analysed during the current study are available from the corresponding author on reasonable request.

## Code availability

The basic HMM scripts are available in https://github.com/predictive-clinicalneuroscience/HMM_Digital_Phenotyping and the imputation script is available in https://github.com/predictive-clinicalneuroscience/Imputation_Digital_Phenotyping

## Supplementary Materials

Supplementary Materials 1: the complete Smartphone Usage and Lifestyle Questionnaire.

