## Supplementary Materials 1: the complete Smartphone Usage and Lifestyle Questionnaire. for "More than one piece of the puzzle: considering non-clinical factors for personalisation in digital phenotyping"

### 4. Intake visit - Smartphone usage questionnaire

| Number | Question | Answers |
|---|---|---|
| 4.1 | Do you always carry your smartphone with you, or have it near you (i.e. in the same room)? | ○ Yes ○ No |
| 4.1.1 | ***If 'Do you always carry your smartphone with you, or have it near you (i.e. in the same room)?' is equal to 'No' answer this question:***<br>When do you not have your smartphone with/near you? (Preferably give specific days, time of day & duration, and if not possible then another indicator, such as 'during work meetings') | |
| 4.2 | Do you ever disable features on your phone? (e.g. location, flight mode, phone on silent) | ○ Yes ○ No |
| 4.2.1 | ***If 'Do you ever disable features on your phone? (e.g. location, flight mode, phone on silent)' is equal to 'Yes' answer this question:***<br>Which features? | |
| 4.2.2 | ***If 'Do you ever disable features on your phone? (e.g. location, flight mode, phone on silent)' is equal to 'Yes' answer this question:***<br>When? (Preferably give specific days, time of day & duration, and if not possible then another indicator, such as 'during work meetings') | |
| 4.3 | Do you ever completely switch off your smartphone? | ○ Yes ○ No |
| 4.3.1 | ***If 'Do you ever completely switch off your smartphone?' is equal to 'Yes' answer this question:***<br>If yes, when? (Preferably give specif ic days, time of day & duration, and if not possible then another indicator, such as 'during work meetings') | |
| 4.4 | Do you use your personal smartphone for work? | ○ Yes ○ No |
| 4.4.1 | ***If 'Do you use your personal smartphone for work?' is equal to 'Yes' answer this question:***<br>If yes, when? (Preferably give specific days, time of day & duration, and if not possible then another indicator, such as 'during work meetings') | |
| 4.5 | Do you feel that you are dependent to some extent on your smartphone? | ○ Yes ○ No |

| | | | |
|---|---|---|---|
| 4.5.1 | ***If 'Do you feel that you are dependent to some extent on your smartphone?' is equal to 'Yes' answer this question:*** To what extent? Please rate from 1 to 5 (1: minor dependence, 3: moderate dependence, 5: extremely dependent) | minor dependence (1) | extremely dependent (5) |
| 4.5.2 | ***If 'Do you feel that you are dependent to some extent on your smartphone?' is equal to 'Yes' answer this question:*** Please give any possible reasons for your dependence (e.g. phone needed for work, to communicate with family) | | |
| 4.6 | Do you feel that you are addicted to some extent to your smartphone? | ○ Yes ○ No | |
| 4.6.1 | ***If 'Do you feel that you are addicted to some extent to your smartphone?' is equal to 'Yes' answer this question:*** To what extent? Please rate from 1 to 5 (1: minor addiction, 3: moderate addiction, 5: extremely addicted) | minor addiction (1) | extremely addicted (5) |
| 4.6.2 | ***If 'Do you feel that you are addicted to some extent to your smartphone?' is equal to 'Yes' answer this question:*** Please give any possible reasons for your addiction (e.g. phone use is a coping mechanism, level of phone use viewed as acceptable amongst peers) | | |
| 4.7 | Do you perceive any barriers to using your smartphone? E.g. any concerns that you have about the privacy of your data, or practical aspects that affect how easily you can operate your phone (difficulties typing, impaired vision, difficulties reading on the phone, etc.) | ○ Yes ○ No | |
| 4.7.1 | ***If 'Do you perceive any barriers to using your smartphone?' is equal to 'Yes' answer this question:*** Please provide a brief explanation of the perceived barrier | | |
| 4.7.2 | ***If 'Do you perceive any barriers to using your smartphone?' is equal to 'Yes' answer this question:*** What phone apps does this affect? (e.g. dexterity issues affecting communication app use; privacy concerns affecting social media app use, Google maps use) | | |
| | Other technology | | |
| 4.8 | Do you use another phone that is not your personal smartphone? | ○ Yes ○ No | |

| | | |
|---|---|---|
| 4.8.1 | ***If 'Do you use another phone that is not your personal smartphone?' is equal to 'Yes' answer this question:*** What kind of phone? (e.g. a work phone) | |
| 4.8.2 | ***If 'Do you use another phone that is not your personal smartphone?' is equal to 'Yes' answer this question:*** Approximately when? (Preferably give specific days, time of day & duration) | |
| 4.9 | Do you use a computer (PC or laptop), tablet, or console for gaming? | ○ Yes ○ No |
| 4.9.1 | ***If 'Do you use a computer (PC or laptop), tablet, or console for gaming?' is equal to 'Yes' answer this question:*** Approximately when? (Preferably give specific days, time of day & duration) | |
| 4.9.2 | ***If 'Do you use a computer (PC or laptop), tablet, or console for gaming?' is equal to 'Yes' answer this question:*** When you are gaming, do you communicate with others through the game? | ○ Yes ○ No |
| 4.9.2.1 | ***If 'When you are gaming, do you communicate with others through the game?' is equal to 'Yes' answer this question:*** For what approximate portion of your game time do you communicate with others through the game? | |
| 4.10 | Do you use a computer (PC or laptop) or tablet for communicating with people? (e.g. WhatsApp desktop or Facebook) | ○ Yes ○ No |
| 4.10.1 | ***If 'Do you use a computer (PC or laptop) or tablet for communicating with people? (e.g. WhatsApp desktop or Facebook)' is equal to 'Yes' answer this question:*** Approximately when? (Preferably give specific days, time of day & duration) | |
| 4.11 | Do you use a computer (PC or laptop) or tablet for social media? | ○ Yes ○ No |
| 4.11.1 | ***If 'Do you use a computer (PC or laptop) or tablet for social media?' is equal to 'Yes' answer this question:*** Approximately when? (Preferably give specific days, time of day & duration) | |

| | | |
|---|---|---|
| 4.12 | Do you use a computer (PC or laptop), TV, or tablet for other entertainment? (e.g. watching YouTube, Netflix) | ◯ Yes ◯ No |
| 4.12.1 | ***If 'Do you use a computer (PC or laptop), TV, or tablet for other entertainment? (e.g. watching YouTube, Netflix)' is equal to 'Yes' answer this question:*** Approximately when? (Preferably give specific days, time of day & duration) | |
| 4.13 | Are there any other technologies you regularly use? (e.g. VR headset) | ◯ Yes ◯ No |
| 4.13.1 | ***If 'Are there any other technologies you regularly use? (e.g. VR headset)' is equal to 'Yes' answer this question:*** Which technologies? | |
| 4.13.2 | ***If 'Are there any other technologies you regularly use? (e.g. VR headset)' is equal to 'Yes' answer this question:*** Approximately when? (Preferably give specific days, time of day & duration) | |
| | Lifestyle questions | |
| 4.14 | What kind of area do you live in? (e.g. rural, city, suburbs) | |
| 4.15 | Are you currently working and/or studying? | ◯ Yes ◯ No |
| 4.15.1 | ***If 'Are you currently working and/or studying?' is equal to 'Yes' answer this question:*** If yes, specify | ☐ working<br>☐ studying |
| 4.15.2 | ***If 'Are you currently working and/or studying?' is equal to 'Yes' answer this question:*** Could you explain your work schedule (including days off, days worked in office, days worked from home, days worked from alternative locations)? | |
| 4.15.1.1 | ***If 'If yes, specify' is equal to 'working' answer this question:*** For your work: specify your work hours and times (e.g. if you work the same hours each workday or work in varying shifts, including night shifts, and when these shifts start/end) | |

| | | |
|---|---|---|
| 4.15.1.2 | ***If 'If yes, specify' is equal to 'studying' answer this question:***<br>For you study: Give any possible details about regular workdays and times | |
| 4.15.3 | ***If 'Are you currently working and/or studying?' is equal to 'Yes' answer this question:***<br>How long does it take you to get to your work/place of study? | |
| 4.16 | Do you have a mostly regular sleep schedule? | ◯ Yes ◯ No |
| 4.16.1 | ***If 'Do you have a mostly regular sleep schedule?' is equal to 'No' answer this question:***<br>Is there a reason for this? (e.g. work - related, health issue) | |
| 4.16.2 | ***If 'Do you have a mostly regular sleep schedule?' is equal to 'Yes' answer this question:***<br>Approximately when do you go to sleep and wake up? | |
| 4.17 | Do you have regular days that are different to most other days? (e.g. getting up early on workdays and then sleeping in on the weekend) | ◯ Yes ◯ No |
| 4.17.1 | ***If 'Do you have regular days that are different to most other days? (e.g. getting up early on workdays and then sleeping in on the weekend)' is equal to 'Yes' answer this question:***<br>What are these days and the corresponding times? | |
| 4.18 | Do you have other regular routines? (e.g. visiting family, sport classes)<br>regular activities that could help us interpret missing data during specific periods throughout the week | ◯ Yes ◯ No |
| 4.18.1 | ***If 'Do you have other regular routines? (e.g. visiting family, sport classes)' is equal to 'Yes' answer this question:***<br>Describe each activity | |

| | |
|---|---|
| 4.18.2 | ***If 'Do you have other regular routines? (e.g. visiting family, sport classes)' is equal to 'Yes' answer this question:***<br>When do you carry out each activity? (Preferably give specific days, time of day & duration) |
| 4.19 | Which modes of transport do you use? |
| 4.20 | Regarding the modes of transport that you use: If you have specific routines (e.g. bus for commuting, walking to the shops), please give details (Preferably give specific days, time of day & duration) |
