## Supplementary Materials 2: additional figures and further methodological information regarding multicollinearity of the mean transition probabilities. for "More than one piece of the puzzle: considering non-clinical factors for personalisation in digital phenotyping"

### Figures

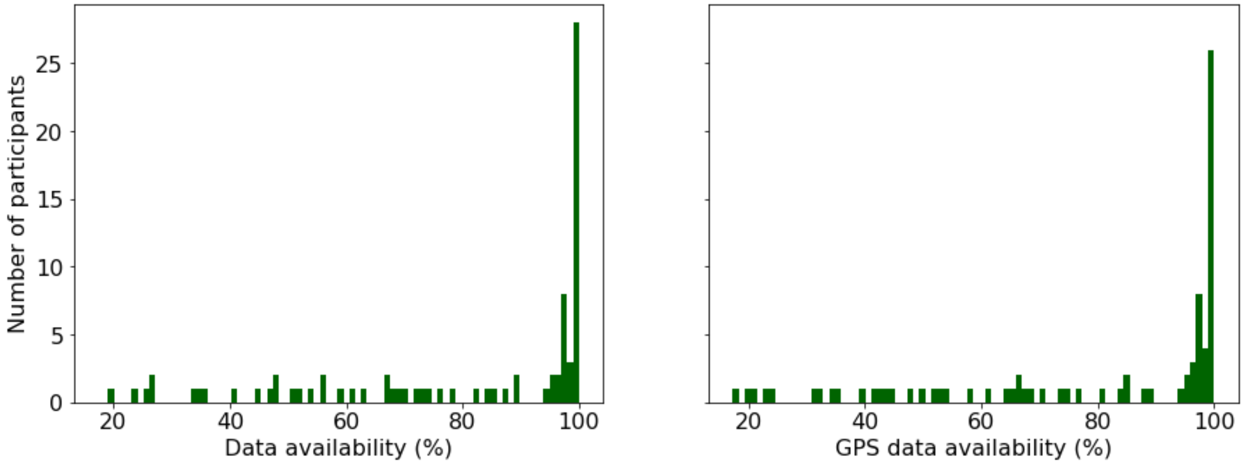


*Figure S1: Histograms of the overall percentage of data available per participant (left) and percentage of GPS data available (right).*

### Methods

Further methodological information regarding multicollinearity of the mean transition probabilities, and the subsequent selection of mean transition probabilities for SULQ outcome prediction, is provided in this section.

As we had multiple classes in the SUQ_area item, we used the multinom function to predict outcomes. However, the multicollinearity of mean transition probabilities caused issues in model estimation, and discrepancies were observed between multinom estimation and glm estimation for binomial outcomes. A subset of mean transition probabilities therefore needed to be selected for more reliable model estimation.

Mean transition probabilities that were near 0 for all participants were removed. All transitions from state 1 had a good range so could be included, but as glm excludes one of these transitions (unlike multinom) due to singularities (all probabilities sum to 1), we checked their correlations to see if any of the transitions was most strongly correlated with the other transitions. The state 1 to state 5 transition was the only transition that had the highest correlation with more than one of the other possible transitions, so this transition was removed as a predictor. Having removed near-0 mean transitional probabilities and the state 1 to 5 transition, we began our collinearity investigations using this equation:

model <- glm(SUQ_addicted ~ state_1to1_prob_mean + state_1to2_prob_mean + state_1to3_prob_mean + state_1to4_prob_mean + state_2to1_prob_mean + state_2to2_prob_mean + state_2to4_prob_mean + state_2to5_prob_mean + state_3to1_prob_mean + state_3to3_prob_mean + state_3to5_prob_mean + state_4to1_prob_mean + state_4to2_prob_mean + state_4to4_prob_mean +

state_4to5_prob_mean + state_5to2_prob_mean + state_5to4_prob_mean + state_5to5_prob_mean + age, family=binomial(link="logit"), data = df_testing)

14 of the transitions were classified by the check_collinearity function as being highly correlated (according to their VIF). state_2to5_prob_mean had the highest VIF. This was therefore dropped from the glm formula and check_collinearity was run again. The remaining transitions from state 2 were now categorised as having low correlations. In this new model, state_4to4_prob_mean had the highest VIF. The process was repeated, with the remaining transitions from state 4 now considered to have low correlation. In the next iteration, state_3to3_prob_mean had the highest VIF and again, after dropping this predictor the remaining transitions from state 3 showed low correlations. Finally, state_5to2_prob_mean had the highest VIF, and after being dropped all remaining predictors showed low correlations. The highest VIF of these predictors was 3.38 (state_4to1_prob_mean). The final list of predictors that were used across all outcomes was therefore: state_1to1_prob_mean, state_1to2_prob_mean, state_1to3_prob_mean, state_1to4_prob_mean, state_2to1_prob_mean, state_2to2_prob_mean, state_2to4_prob_mean, state_3to1_prob_mean, state_3to5_prob_mean, state_4to1_prob_mean, state_4to2_prob_mean, state_4to5_prob_mean, state_5to4_prob_mean, state_5to5_prob_mean, age
